# Human genomic regions of systemic interindividual epigenetic variation are implicated in neurodevelopmental and metabolic disorders

**DOI:** 10.1101/2025.02.10.25322021

**Authors:** Wen-Jou Chang, Uditha Maduranga, Chathura J. Gunasekara, Alan Yang, Matthew Hirschtritt, Katrina Rodriguez, Cristian Coarfa, Goo Jun, Craig L. Hanis, James M. Flanagan, Ivan P. Gorlov, Christopher I. Amos, Joseph L. Wiemels, Catherine A. Schaefer, Ezra S. Susser, Robert A. Waterland

## Abstract

Epigenome-wide association studies (EWAS) profile DNA methylation across the human genome to identify associations with diseases and exposures. Most employ Illumina methylation arrays; this platform, however, under-samples interindividual epigenetic variation. The systemic and stable nature of epigenetic variation at correlated regions of systemic interindividual variation (CoRSIVs) should be advantageous to EWAS. Here, we analyze 2,203 published EWAS to determine whether Illumina probes within CoRSIVs are over-represented in the literature. Enrichment of CoRSIV-overlapping probes was observed for most classes of disease, particularly for neurodevelopmental disorders and type 2 diabetes, indicating an opportunity to improve the power of EWAS by over 200- and over 100-fold, respectively. EWAS targeting all known CoRSIVs should accelerate discovery of associations between individual epigenetic variation and risk of disease.

## Main Text

Genomic studies including genome-wide association studies (GWAS) and whole-genome and exome sequencing have expanded our understanding of complex diseases by enabling identification and characterization of genetic variants associated with specific diseases and traits (*1–6*). Despite this success, for most common diseases the genetic variants identified by genomic studies explain only a fraction of the risk (*7*), suggesting a role for individual epigenetic variation in disease susceptibility (*8, 9*). For over a decade, epigenome-wide association studies (EWAS) have focused on profiling CpG methylation at the genome scale to uncover links between epigenetic variation and diseases or phenotypes (*10, 11*). Although thousands of studies have been conducted, there are few robust and replicable associations. Recent reviews summarizing EWAS conducted using the Illumina methylation platform (*12–14*) reveal poor replicability across studies encompassing a range of diseases and outcomes, leading some to question if epigenetic variation is indeed a major factor in disease risk (*15*).

An important consideration, however, is that the Illumina methylation platform, the workhorse of EWAS, was not designed for detection of population level associations. The success of genomic studies, in particular GWAS, was built upon the HapMap (*16*) and 1000 Genomes (*17*) projects, which identified sites of common sequence variation in the human genome so they could be assayed at the population level. The EWAS era, however, was launched without an analogous project to map out human genomic regions of interindividual epigenetic variation. Instead, investigators adopted the Illumina methylation array platform, principally the HM450 and then the EPIC 850 array, which were designed to detect tissue- and gene-level DNA methylation profiles. As these arrays were designed without knowledge of interindividual epigenetic variation (*18, 19*), ∼90% of the probes (depending on the specific criteria) target CpG sites that show negligible interindividual variation in normal somatic tissues (*20–23*), compromising their ability to detect associations. Indeed, the low interindividual variation of most probes on the Illumina methylation platform is associated with low technical reliability (*20, 21*) and poor stability in test-retest experiments over time (*22, 23*).

Whereas the need for interindividual variation is common to GWAS and EWAS, factors unique to EWAS also warrant consideration (*9*). Unlike DNA sequence, DNA methylation is largely cell type-specific (*24*). Nonetheless, most EWAS conducted so far have profiled methylation in blood DNA, even when attempting to draw associations with diseases whose pathophysiology is putatively unrelated to leukocyte gene expression, such as neurological disorders or type 2 diabetes. Also unlike DNA sequence, methylation at some CpG sites is potentially dynamic, such that disease progression can induce epigenetic changes (*25, 26*). To avoid such reverse causality, identifying variants with long-term temporal stability is advantageous for EWAS.

Accordingly, ideal genomic regions to target in population epigenetic studies would show substantial interindividual variation in DNA methylation that is both systemic (i.e. not cell type specific) and stable over time. To identify such regions, we previously conducted deep unbiased DNA methylation profiling in three embryonic germ layer lineages of each of ten human donors, identifying ∼10,000 correlated regions of systemic interindividual epigenetic variation (CoRSIVs) (*27*). A subsequent study of 188 individuals (*28*) validated systemic interindividual variation (SIV) at CoRSIVs and also identified over 70 times more genetic influence on methylation (methylation quantitative trait loci – mQTL) than had been detected by other studies, mostly based on the Illumina platform. Just as for detection of mQTL, interindividual variation in DNA methylation is essential for detection of associations with disease and environmental exposures. We therefore postulated that, relative to most probes on the Illumina array platform, those within CoRSIVs are more likely to be reported in EWAS. Here, we describe an automated text-mining analysis testing this hypothesis.

### Across 2,203 published EWAS, Illumina probe ID occurrences link to biology

Medical Subject Headings (MeSH) comprise a defined, hierarchically organized vocabulary used in indexing articles in PubMed. Based on disease categories in a previous study (*28*), and the MeSH hierarchy under “diseases” [MeSH], we selected 11 disease categories for analysis: cancer, cardiovascular, digestive, endocrine, hematological, immune, metabolic, neurological (including both nervous system and mental disorders), obesity, respiratory, and urogenital (Fig. 1A). From the full-text and supplementary tables of 2,203 publications under these 11 categories (Fig. 1A, B, table S1), we extracted 234,586 unique HM450 / EPIC probe IDs (which we refer to as ‘probes’). Across each of 11 disease categories, the number of publications ranged from 85 (hematological) to 1,085 (cancer) (Fig. 1C). We chose not to use existing EWAS databases such as the EWAS Atlas (*29*) or EWAS catalog (*30*) because these manually curated databases are inherently limited in scope. The largest, EWAS Atlas, includes only ∼1,000 studies, fewer than half as many as we evaluated (fig. S1).

**Fig. 1.**
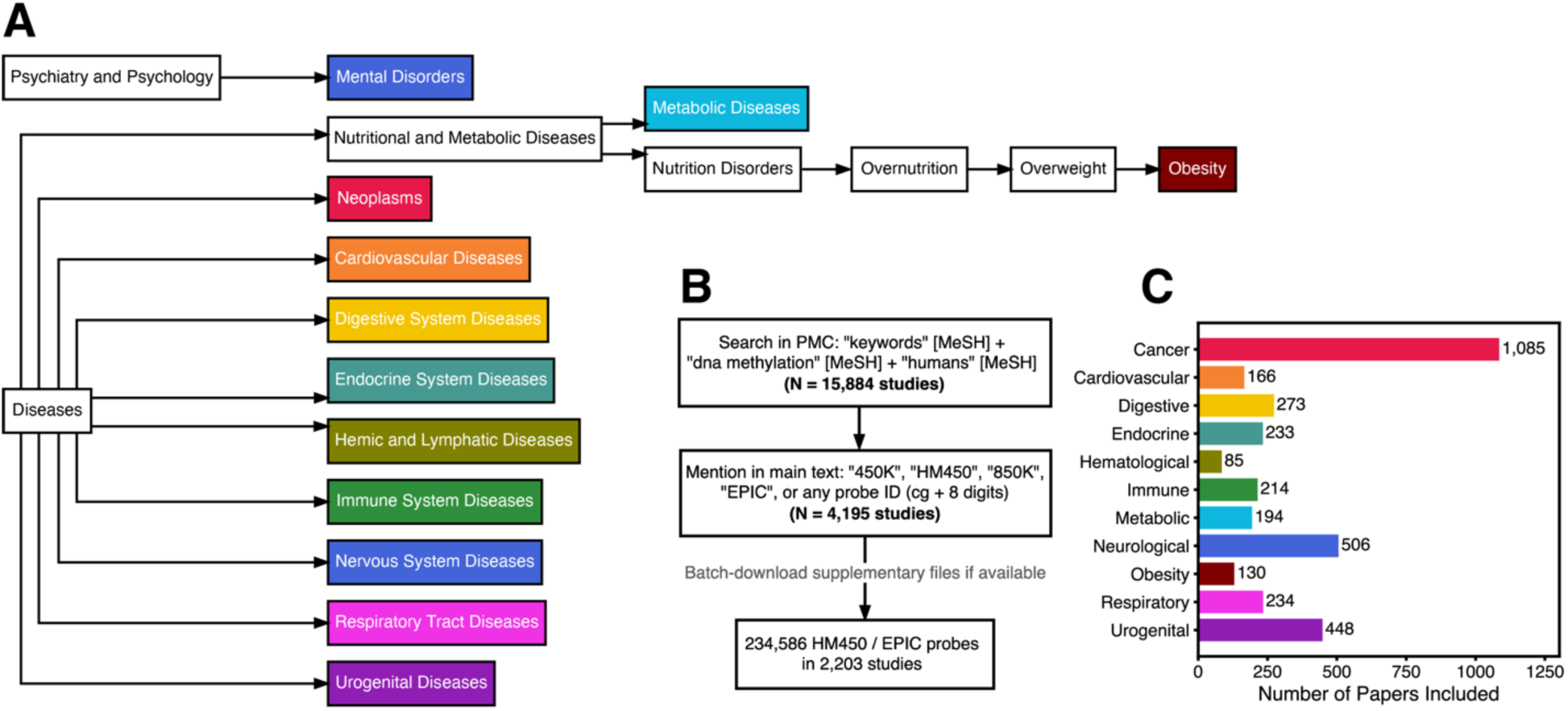
Workflow for mining Illumina probe IDs from the EWAS literature. (**A**) Hierarchical tree of Medical Subject Headings (MeSH) terms included in the analysis. (**B**) Workflow for paper selection and probe extraction process. (**C**) Number of studies included within each disease category. Category labels are abbreviations of the MeSH terms in panel A, except for “Neurological”, which combines MeSH terms “Mental Disorders” and “Nervous System Diseases”.

Our analysis is predicated on the assumption that probes reported in the literature in the context of different MESH terms reflect, to some extent, associations with pathophysiology of specific diseases. To test this, taking cancer as an example, we observed that as we increase the number of papers in which a specific probe must be reported, the number of probes meeting the threshold decays exponentially (Fig. 2A). Considering the 18,132 probes reported in at least one paper in the context of cancer (but not in other disease categories), we explored the associated genes by gene-set enrichment analysis using DisGeNET. Although the top 10 disease terms (by adjusted P value) included several cancer-related terms, these were barely significant (Fig. 2B). When we restricted the analysis to the 13,113 probes reported in two or more studies in the context of cancer, the top ten disease terms were highly significant and all related to cancer (Fig. 2B). This indicates that probes reported in at least two papers in the context of the same disease are more reliably linked to pathophysiology.

**Fig. 2.**
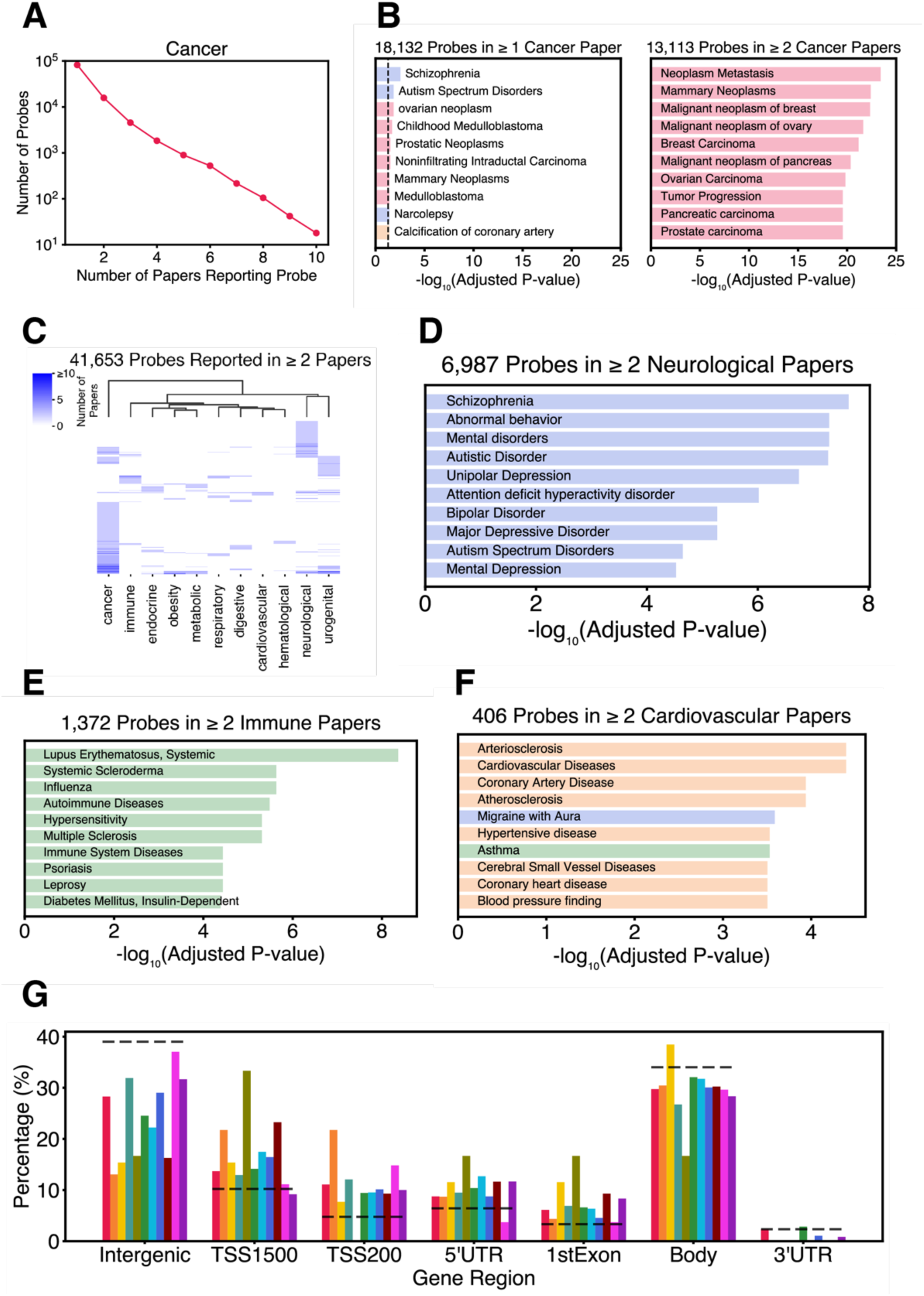
Probe ID occurrences in the literature map to biology. (**A**) The decay plot for cancer illustrates that as we increase the number of papers in which a specific probe must be reported, the number of probes meeting the threshold decays exponentially. (**B**) Gene set enrichment analysis (GSEA) of genes annotated to the 18,132 probes cited in at least one cancer-related paper (and unique to cancer). The top ten terms (by adjusted P value) are moderately significant and include some unrelated to cancer. Restricting the analysis to the 13,113 probes reported in two or more cancer-related studies yields highly significant terms, all related to cancer. (Terms are shaded as in Fig. 1C.) (**C**) Only 41,653 probes were reported in two or more papers within at least one of the 11 disease categories. Unsupervised hierarchical clustering of these reveals distinct clusters of probes associated with cancer and neurological disorders, and overlap in related categories such as metabolic and obesity. (**D-F**) GSEA of probes reported in at least two papers within each of the (**D**) neurological, (**E**) immune, and (**F**) cardiovascular categories identifies highly significant terms primarily linked to the respective classes of disease. (**G**) Compared to all CoRSIVs covered on the Illumina platform (dotted lines), those reported in at least 2 papers within each of several disease categories tend to locate near the transcription start site (TSS) of genes (P < 10^-10^) and not in intergenic regions (P = 0.006). Each colored bar represents CoRSIVs reported in ≥ 2 papers within each disease category; color-coding follows Figure 1C. Dashed line represents all 1,607 CoRSIVs covered on the Illumina platform.

Only 41,653 probes were reported in at least 2 papers within the same disease category (table S2). Unsupervised hierarchical clustering of these (Fig. 2C) identified both distinct clusters of probes uniquely associated with specific categories of disease (such as cancer and neurological disorders) and extensive overlap in some related categories such as metabolic and obesity. Notably, excluding the 13,113 probes reported only in the context of cancer, only 28,460 probes were reported in at least two papers in the context of any of the other diseases. Similar to the results for cancer (Fig. 2B), gene-set enrichment analysis considering genes associated with probes uniquely reported in two or more studies in the context of neurological, immune, or cardiovascular disorders detected highly significant disease terms primarily associated with the respective categories (Fig. 2D – F; table S3).

We compiled curated systemic interindividual variation (SIV) loci from several sources including putative metastable epialleles (*31*), Illumina probes identified as exhibiting epigenetic supersimilarity or systemic interindividual variation (*32*), and the 9,926 human CoRSIVs (*27*). Although some of these loci were identified prior to introduction of the CoRSIV terminology, we consider them all CoRSIVs. Of these 10,388 regions, only 1,607 (15.5%) are covered by at least one probe on the Illumina HM450 or EPIC array (table S4). Within these 1,607 regions are 3,517 probes, which we term CoRSIV probes (average 2.2 probes/CoRSIV); table S5 annotates these relative to proximal genes.

We next asked whether CoRSIVs reported in the literature tend to be associated with specific genic regions. Compared to all 1,607 CoRSIVs covered on the Illumina array, those reported in at least 2 papers in the context of one of the 11 disease categories were about twice as likely to be within 200 bp of a transcription start site (TSS200) of a gene (chi-square P < 10^-10^) and half as likely to be intergenic (chi-square P = 0.006) (Fig. 2G). Together, these results support the plausibility of our analysis by indicating that occurrences of probe IDs in the literature associate with disease-specific pathophysiology and reflect the established role of promoter methylation in transcriptional regulation (*33*).

### CoRSIV probes are overrepresented in most disease categories

Considering the relatively high CpG density of CoRSIVs (≥5 CpGs per 200 bp), the full set of Illumina probes is not an appropriate background set against which to evaluate enrichment of CoRSIV probes. We therefore modeled expected probe occurrences based on 10 sets of control regions; within each of the 10 sets, a control region was matched to each CoRSIV based on a range of parameters including chromosome, region size, and number of overlapping Illumina probes (see Methods). This yielded 35,170 control probes (table S6) within 16,070 control regions (table S7). We averaged across the ten sets of control regions to establish a stable baseline from which to evaluate enrichment. We noted that, for most classes of disease, as we increased the threshold for the number of papers in which a specific probe must be reported (i.e., increasing reliability), the percentage of those probes overlapping CoRSIVs increased markedly (Fig. 3A – F); no such increase was observed among the control probes (gray lines).

**Fig. 3.**
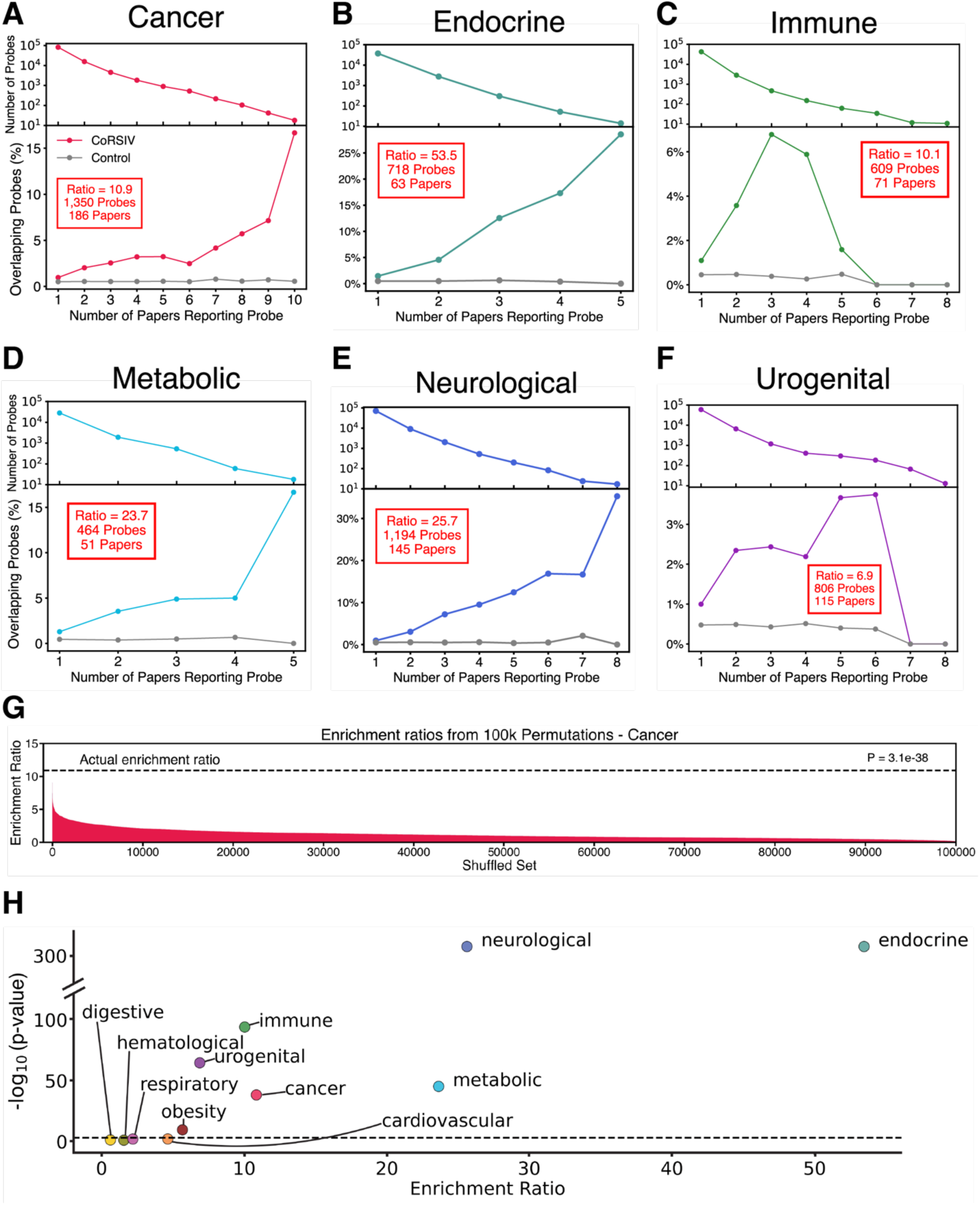
In most disease categories, probes reported in the literature are enriched for overlap with CoRSIVs. (**A-F**) Stacked decay-enrichment plots for the cancer, endocrine, immune, metabolic, neurological, and urogenital categories, respectively. As individual probes are required to be reported in more papers, the number of probes meeting the threshold decays (top panels) while the percent overlap with CoRSIVs increases (lower panels). Insets (red boxes) report the enrichment ratio and number of CoRSIV probes and papers driving each enrichment. (**G**) Results of permutation testing (100,000 iterations) for the cancer category. (**H**) Summary of results across all 11 categories.

Within each disease category, we assessed enrichment of CoRSIV relative to control probes by calculating the ratio of the weighted sum of CoRSIV probe occurrences to the average weighted sum of control probe occurrences, with weights based on the number of publications reporting each probe:

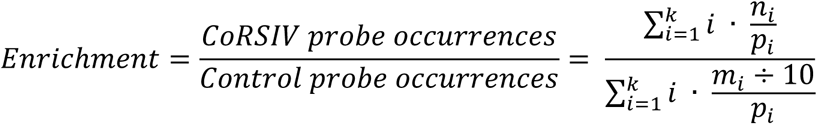

where ***k*** is the highest number of papers that each report at least 10 identical probe IDs, ***n_i_*** is the number of CoRSIV probes reported in exactly ***i*** papers, ***p_i_*** is the total number of probes reported in exactly ***i*** papers, and ***m_i_*** is the number of control probes reported in exactly ***i*** papers (across 10 control sets). We then determined statistical significance by permutation testing (Fig. 3G and fig. S2). An annotated example of an enrichment ratio calculation (fig. S3) shows that high enrichment ratios are largely driven by CoRSIV probes reported in a high number of papers.

We found statistically significant enrichment of CoRSIV probes in 7 of the 11 categories: cancer, endocrine, immune, metabolic, neurological, obesity, and urogenital (Fig. 3A – 3F; fig. S4; table S8). Results for all categories are summarized in Fig. 3H. The strongest enrichment ratios were found within the endocrine (53.5-fold), neurological (25.7-fold), and metabolic categories (23.7-fold); cancer and immune disorders yielded over 10-fold enrichments. Remarkably, we observed these strong overrepresentations of CoRSIV probes despite only 15.5% of known CoRSIVs being represented on the Illumina platform. Hence, to estimate the power advantage of profiling all known CoRSIVs compared to using the Illumina platform, these enrichments can be multiplied by 6.5 (i.e. 1/0.155). EWAS profiling all known CoRSIVs would therefore be expected to have 345-, 166-, and 153-fold greater power to detect associations with endocrine, neurological, and metabolic disorders, respectively (fig. S5; table S8). Even for diseases showing modest enrichment, like cancer, urogenital disorders and obesity, CoRSIV-focused EWAS promise remarkable 70-, 45- and 37-fold increases in power, respectively.

### High-resolution analyses provide insights into category-specific enrichments

Each of the MeSH categories we evaluated comprises dozens of different diseases and disorders. For several categories with ample numbers of papers, therefore, we utilized the MeSH hierarchy to conduct enrichment analyses at higher resolution. For example, within the neurological category (Fig. 4A; table S9), CoRSIVs were particularly enriched in the context of neurodevelopmental disorders (34.6-fold; P = 1.1 x 10^-187^; Fig. 4B), including intellectual disability (22.5-fold; P = 5.3 x 10^-71^; Fig. 4C), attention deficit disorder with hyperactivity (54.7-fold; P < 2.2 x 10^-308^; Fig. 4D), autism spectrum disorder (21.1-fold; P = 4.0 x 10^-108^; fig. S6A), and schizophrenia (12.7-fold; P = 1.0 x 10^-97^; fig. S6B). (Although not listed within “neurodevelopmental disorders” in the MeSH hierarchy, many consider schizophrenia a neurodevelopmental disorder (*34*).) These enrichments have not been multiplied by the 6.5 scaling factor. Remarkably, over half of the papers within the neurodevelopmental disorders and ADHD categories, and two-thirds of those under intellectual disability report at least one CoRSIV probe. In striking contrast, and despite comparable numbers of studies, neurodegenerative disorders including dementia, tauopathies, and Alzheimer’s disease showed no enrichment (Fig. 4A). In addition to neurodevelopmental disorders, our analysis (Fig. 4A) detected an enrichment of CoRSIV probes in studies associated with nervous system diseases including nervous system neoplasms (22.3-fold; P < 2.2 x 10^-308^; fig. S6C) and multiple sclerosis (23.5-fold; P = 8.4 x 10^-291^; fig. S6D).

**Fig. 4.**
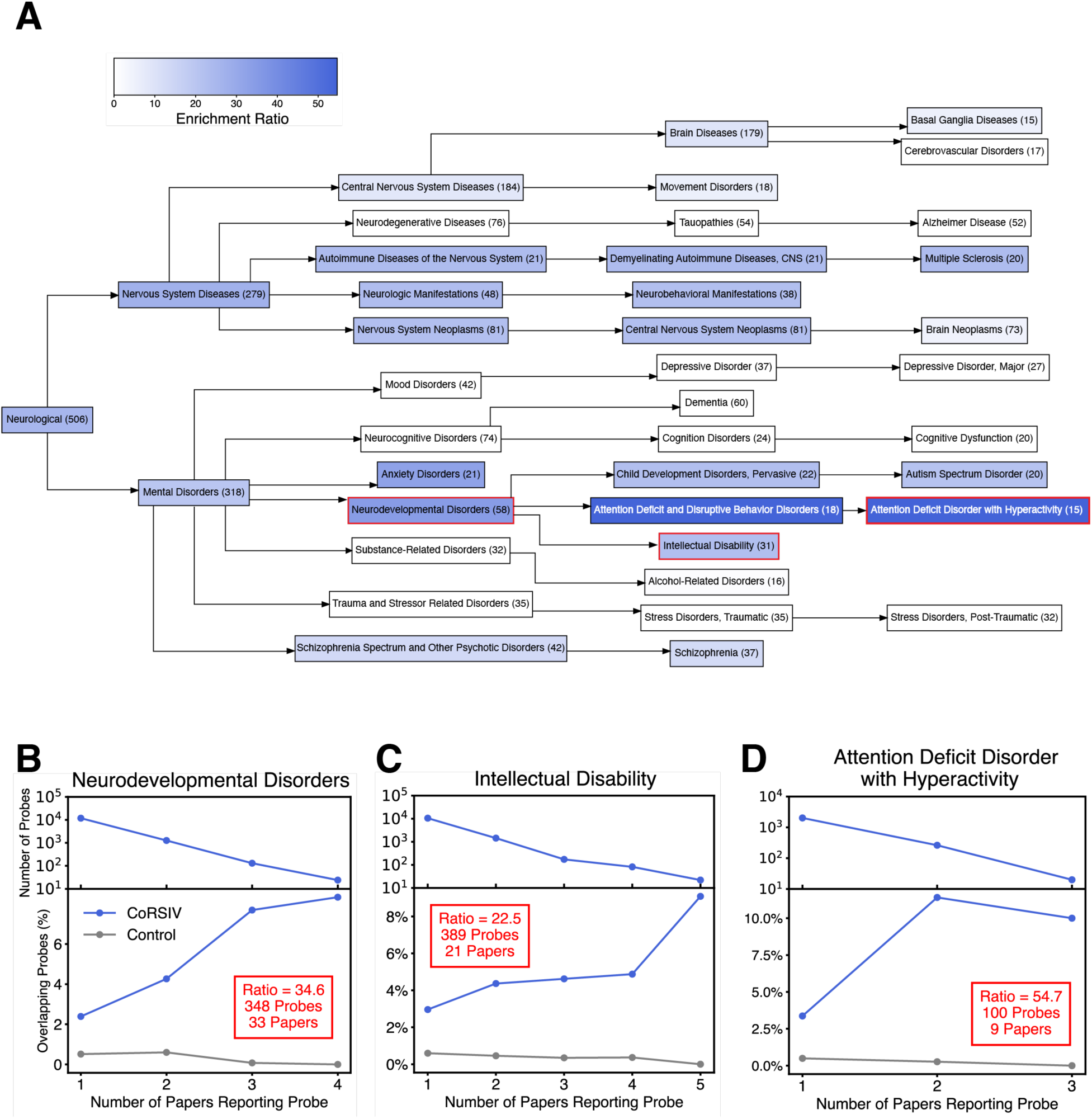
CoRSIVs are associated with neurodevelopmental but not neurodegenerative disorders. (**A**) Term-specific results for the MeSH hierarchy encompassing the neurological category. Statistically significant enrichments are indicated by blue shading. Numbers indicate the total number of papers analyzed within each MeSH term. Only categories showing (i) statistically significant enrichment, and (ii) including at least one CoRSIV probe reported in at least two papers are colored. (**B-D**) Stacked decay and enrichment plots for neurodevelopmental disorders, intellectual disability, and attention deficit disorder with hyperactivity, respectively.

Within our text-mining approach we were unable to develop a reliable automated method to focus only on statistically significant probe occurrences. In the context of this sub-category analysis, therefore, we tested whether restricting the analysis to statistically significant probes yields comparable results. We manually curated all 15 papers annotated to the “Attention Deficit Disorder with Hyperactivity” MeSH Term and tabulated nominally significant probes associated with ADHD or environmental exposures leading to ADHD. Whereas 1,555 probe instances were common to both approaches (fig. S7A), 1,399 were unique to the manual approach, as the automated approach was blind to tables not included in the PMC Open Access Subset. Of the 1,054 probe instances unique to the automated approach, ∼60% were excluded from the manual approach because they were not directly relevant to ADHD outcomes and ∼40% were because they were not statistically significant. Overall, the manual approach, yielded an enrichment ratio of 34.6 (table S10; P = 3.0 x 10^-276^; fig. S7B), comparable to the 54.7 ratio obtained by the automated approach.

Enrichment analysis within the metabolic MeSH category (fig. S8A; table S11) indicated that its enrichment was largely driven by glucose metabolism disorders (45.7-fold enrichment; P = 7.8 x 10^-170^; fig. S8B), specifically type 2 diabetes (15.8-fold; P = 2.0 x 10^-32^; fig. S8C) and gestational diabetes (9.3-fold; P = 8.0 x 10^-27^; fig. S8D), as well as lipid metabolism disorders (18.1-fold; P < <2.2 x 10^-308^). With the caveat that there were a relatively small number of studies, no enrichment was observed for type 1 diabetes. Exploring the endocrine MeSH category (fig. S9; table S12) indicated that, like metabolism, its exceptionally strong enrichment was primarily driven by type 2 diabetes and gestational diabetes.

Lastly, breaking down cancer, the MeSH category with the largest number of papers (fig. S10A; table S13), indicated a high degree of heterogeneity. Whereas cancer showed an 11.2-fold over-representation CoRSIV probes (Fig. 3A), most cancer subtypes showed no enrichment. The most striking exception is the 58.1-fold enrichment for prostate cancer (P = 1.2 x 10^-87^; fig. S10B); nervous system neoplasms (22.3-fold; P < 2.2 x 10^-308^; fig. S10C) and neuroendocrine tumors (5.4-fold; P = 2.7 x 10^-14^; fig. S10D) also showed enrichment. MeSH hierarchies and diseases showing significant CoRSIV enrichment in the context of immune and urogenital studies are shown in fig. S11 and table S14, S15. Additional details on studies contributing to CoRSIV enrichment at the specific disease level are provided in tables S16 – S21.

### Stable and Systemic Interindividual Variation Explains the Advantage of CoRSIVs

Wishing to gain insights into why CoRSIV-overlapping probes are so strongly over-represented in the EWAS literature, we analyzed data on long-term stability of DNA methylation in peripheral blood of 92 individuals each profiled twice using the Illumina HM450 methylation array, across an average interval of six years (*35*). We quantified stability of probe-level methylation using the intraclass correlation coefficient (ICC), and interindividual variation using the 2^nd^ to the 98^th^ percentile of interindividual range (IIR_2-98%_) (*36*) (table S22) (see Methods). Probes with an ICC > 0.5 are considered moderately stable (*35*). Evaluating the 2,111 CoRSIV probes and 412,269 non-CoRSIV probes with data available on both ICC and IIR_2-98%_ (Fig. 5A) indicated that long-term stability was strongly associated with interindividual range. CoRSIV probes generally exhibited substantial interindividual variation (median IRR_2-98%_ = 0.22), much greater than that of non-CoRSIV probes (median IRR_2-98%_ = 0.06). CoRSIV probes were also highly stable (median ICC = 0.82), whereas long-term stability of non-CoRSIV probes was generally poor (median ICC = 0.16). These dramatic differences between CoRSIV and non-CoRSIV probes were also observed among probes associated with different classes of disease (fig. S12).

**Fig. 5.**
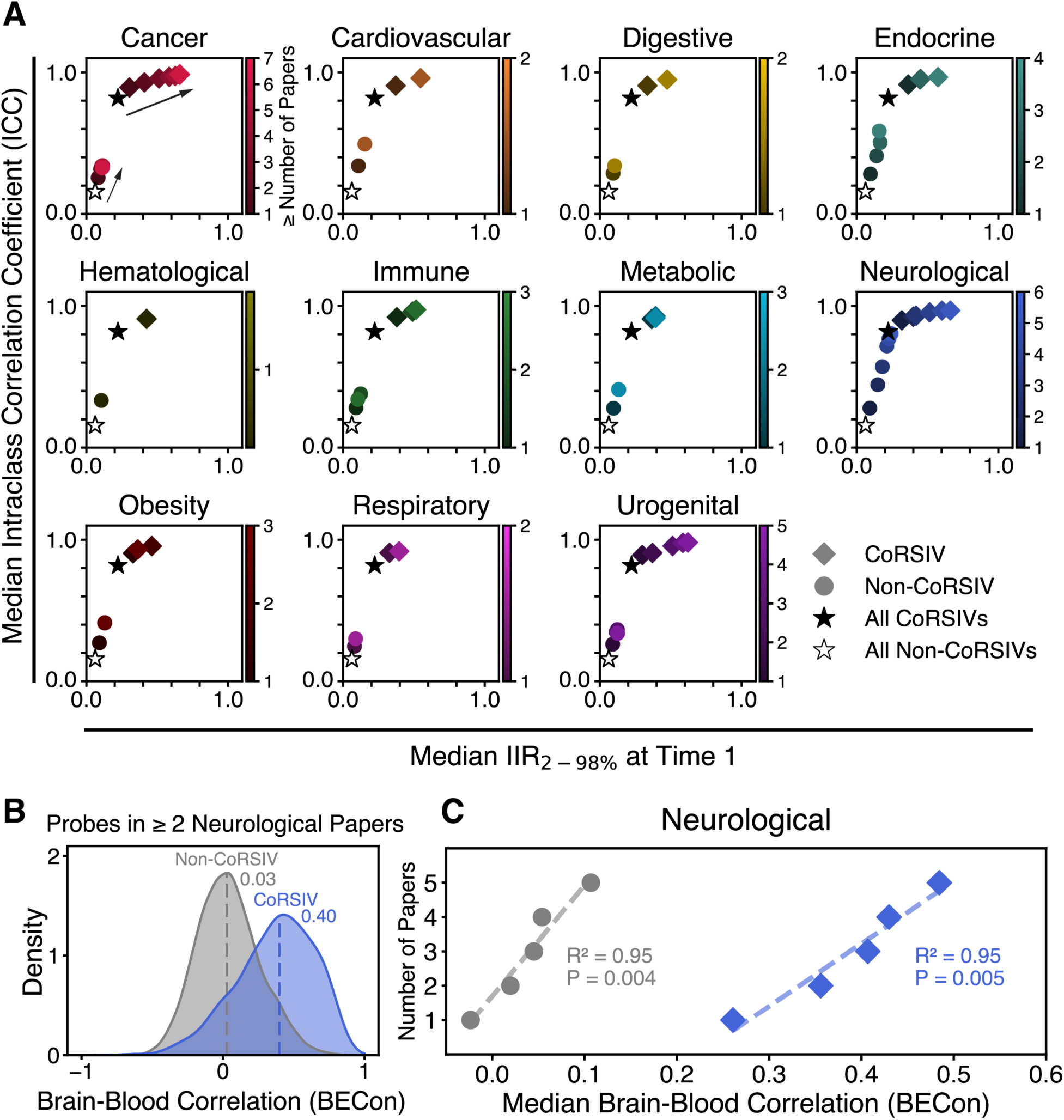
Stable, systemic interindividual variation appears to explain CoRSIVs’ advantage. (**A**) Six-year test-retest stability data based on the HM450 platform (*35*) show that long-term stability (intraclass correlation coefficient, ICC) is dependent on interindividual range (IIR_2-98%_, see Methods). The median ICC of the 2,111 CoRSIV-overlapping probes (filled star) is much higher than that of the 412,269 non-CoRSIV probes (hollow star). Category-specific sensitivity analyses (shading) show that, for CoRSIV probes, being reported in more papers is associated with increasing interindividual range (rightward trend indicated by arrow). Only data points representing at least 15 probes are shown. (**B**) Analysis of probe-level brain vs. blood correlation data (BECon) (*37*) shows that, of those reported in at least two papers within the neurological category, CoRSIV probes (blue) show higher brain-blood correlation than non-CoRSIV probes (grey). Dotted lines indicate median values. (**C**) Sensitivity analysis; being reported in more neurological papers is associated with increasing BECon score, for both CoRSIV and non-CoRSIV probes. Only data points representing at least 15 probes are shown.

In sensitivity analyses, within each MeSH category we asked whether interindividual range and stability of DNA methylation are related to the strength of the associations, based on the numbers of papers reporting a specific probe. For CoRSIV probes, being reported in more papers was associated with increasing interindividual range (Fig. 5A), particularly in the cancer, neurological, and urogenital categories (P = 0.001, 1.2 x 10^-4^ and 0.003 respectively) and nearly so for endocrine (P = 0.05). For non-CoRSIV probes, on the other hand, no probe subsets showed median interindividual range greater than 0.2; rather, being reported in more papers was most strongly associated with increasing stability of methylation, most obviously within the endocrine and neurological categories (Fig. 5A). These results indicate that interindividual variation that is substantial and stable increases the likelihood that a probe will be reported in the literature.

Next, to test whether the systemic nature of interindividual variation at CoRSIVs is also factor we queried the BECon database (*37*), which provides probe-level data on correlations of DNA methylation between blood and four different brain regions. The median blood-brain correlation (BECon score) was substantially higher for CoRSIV than non-CoRSIV probes, not only within the neurological category (0.40 and 0.03 respectively) (Fig. 5B) but also within the others (fig. S13A). A sensitivity analysis within the neurological category showed that, for both CoRSIV and non-CoRSIV probes, being reported in more papers was associated with increasing brain-blood concordance (Fig. 5C). In the other disease categories, significant associations for CoRSIV probes were not detected (fig. S13B). This is consistent with the blood-brain basis of the BECon score. Together, these analyses indicate that the over-representation of CoRSIV probes in the EWAS literature is attributable to the substantial, systemic, and stable interindividual variation in DNA methylation at the targeted CpG sites.

## Discussion

In this study, we mined text from 2,203 published studies using the Illumina methylation array platform, finding that the 3,517 Illumina methylation array probes overlapping CoRSIVs are reported in the literature much more than would be expected by chance. Of 11 disease categories analyzed, 7 showed statistically significant enrichment. Moreover, sensitivity analyses indicated that, relative to other probes on the Illumina methylation platform, the considerable power advantage of CoRSIV CpGs is related to their interindividual variation, which is substantial, systemic, and stable.

We are not aware of any previous studies that have taken a similar approach to gain insights into the thousands of EWAS that have been reported in the last 12 years using the Illumina methylation platform. Previous studies (*38, 39*) utilized text-mining approaches to study disease-gene associations in the published literature but, to our knowledge, none has been reported in the context of DNA methylation. Additionally, previous text-mining studies have primarily focused on constructing databases of disease-gene relationships (*40*). In general, Illumina probe IDs reported in EWAS are identified based on statistical tests and can be considered as distinct data elements. Our study establishes a precedent for utilizing probe occurrences in the literature to assess the enrichment of specific genomic regions in EWAS, providing a novel framework for integrating EWAS results across multiple studies. The validity of this approach is supported by our gene-set enrichment analyses. Simply by tabulating probes reported in two or more publications within each of several disease categories and evaluating the associated genes, we detected highly significant enrichments for relevant disease terms.

It is interesting to consider why some classes of disease are strongly associated with CoRSIVs, and others not. We detected the strongest enrichments for CoRSIVs in the context of neurological disorders and metabolic disease (particularly type 2 diabetes). The enrichment for neurological disorders is not surprising, given that nearly half of CoRSIV genes are associated with nervous system diseases or mental disorders (*27*). The strong association with metabolic disorders is also consistent with our previous findings (*28*); analysis of the GWAS catalog showed that, among several different classes of disease, single nucleotide variants associated with metabolic disease are most strongly enriched for mQTL at CoRSIVs. The enrichment observed in the metabolic category was primarily driven by type 2 diabetes and gestational diabetes, with no enrichment under type 1 diabetes. This finding aligns with the established role of gestational diabetes as a risk factor for subsequent type 2 diabetes (*41*), whereas type 1 diabetes follows a distinct etiology.

Inspection of the results in the context of neurological disorders yielded the important insight that the enrichments are limited to neurodevelopmental and not neurodegenerative disorders. This aligns with our previous finding that the developmental establishment of methylation at CoRSIVs occurs in the preimplantation embryo, under the influence of both genetic variation (*28*) and periconceptional environment (*27, 31, 42–44*). The association of CoRSIVs with neurodevelopmental disorders could be interpreted in two ways. First, considering their systemic nature, interindividual variation in methylation at CoRSIVs could influence brain development, directly contributing to the risk of neurodevelopmental disorders. Alternatively, associations of CoRSIV methylation with neurodevelopmental disorders may arise if CoRSIV methylation is a biomarker for environmental conditions during prenatal development that affect both epigenetic development in the early embryo and subsequent neurodevelopment. Interindividual variation in CoRSIV methylation may even represent an evolutionary mechanism for promoting neurodiversity and adaptability in response to changing cultural and environmental demands (*6, 45, 46*), conferring population-level benefits while increasing individual risk for neurodevelopmental disorders.

An epigenetic basis for neurodevelopmental disorders and type 2 diabetes is consistent with the developmental origins paradigm, which suggests that during critical ontogenic periods, environmental factors such as nutrition alter developmental pathways, thus affecting lifelong susceptibility to a wide range of chronic diseases (*8, 9*). Extensive evidence from humans and animal models indicates that the risks of neurodevelopmental disorders and type 2 diabetes are strongly influenced by environment during development. The most compelling human data derive from long-term follow up studies of individuals who were exposed to famine during early development. For example, multiple studies in independent cohorts show that early gestational exposure to famine is associated with a doubling of risk for schizophrenia (*47, 48*). A recent analysis of survivors of the Ukraine famine of 1932-33 found that exposure to famine during early gestation likewise results in a two-fold increased risk for type 2 diabetes in adulthood (*49*). Notably, in both examples, the period of susceptibility (or critical window) during early gestation is consistent with the early embryonic period when CoRSIV methylation is established.

On the other hand, except for prostatic and nervous system neoplasms, despite being the disease most studied from an epigenetic perspective, CoRSIV probes were generally not over-represented in cancer. There are several possible reasons for this. First, unlike other classes of disease, for which investigators are often limited to profiling DNA methylation in blood, many cancer studies compare DNA methylation in tumor and adjacent normal tissue. In this context, the systemic nature of variation at CoRSIVs offers no advantage; likewise for studies profiling blood methylation in the context of hematological disorders. Moreover, the original design of the Illumina array prioritized CpG sites with aberrant methylation in tumors, often a consequence rather than a cause of tumorigenesis. Nonetheless, prospective studies assessing DNA methylation in peripheral blood have shown that baseline measurements of CoRSIV methylation in peripheral blood are associated with the risk of several types of cancer (*32, 50*).

Strengths of our study include the extremely large and systematically curated data set we exploited, comprising thousands of published studies and hundreds of thousands of data points (probe occurrences), and our novel approach of analyzing occurrences of Illumina probe IDs to gain biological insights. These factors enabled the detection of strong and highly significant disease-specific associations, even for some disease categories comprising relatively small numbers of papers. Also, of the diseases found to show significant enrichment for CoRSIVs, the ∼100-fold power advantage we estimated relative to studies using the Illumina platform is consistent with the 72-fold power advantage we reported earlier in the context of detecting mQTL (*28*). Further, our sensitivity analyses using independent data sets provided an explanation for the outsized power advantage of CoRSIVs relative to the other probes on the Illumina platform.

Nonetheless, our study is not without limitations. Despite initial efforts to employ large language models, our automated approach was unable to reliably determine the tissue analyzed in each study. Although most EWAS are based on blood, some profile DNA methylation within a tissue more relevant to the disease of interest, in which case the systemic nature of CoRSIVs is not an advantage. Cancer is one example of this, as described above. Whereas our analysis was conducted at the probe level, many EWAS identify differentially methylated regions (DMRs) comprising multiple CpG sites operating in a coordinated fashion. Due to difficulties determining the specific genome build used and inconsistencies in coordinate formatting, however, we were unable to develop an automated approach to assess overlap of DMRs with CoRSIVs. This is unfortunate because, as CoRSIVs are themselves coordinated regions of variable methylation, DMR-level analyses would be expected to show even stronger enrichment for CoRSIVs than probe-level analyses. Our analysis focused exclusively on EWAS conducted using the Illumina platform. However, CoRSIVs have been identified as top hits in studies utilizing other profiling approaches, such as whole-genome bisulfite sequencing in the context of autism spectrum disorder (*51, 52*) and reduced-representation bisulfite sequencing in the offspring of obese mothers (*53*). Our study is based on the rationale that reliability of a specific probe being associated with a disease is based on reporting frequency, rather than P values determined in each study. We were unable to develop a reliable automated approach for linking P values to probe occurrences. Our manually curated analysis focusing on nominally significant probes reported in the context of ADHD, however, yielded a highly significant enrichment comparable to that obtained using the automated approach, supporting the validity of our results. Nonetheless, the ability to analyze additional information from each study, such as sample sizes and P values, would likely be advantageous.

## Conclusion

Our findings have important implications for the fields of epigenetic epidemiology and population epigenetics. The finding that neurodevelopmental disorders are enriched for CoRSIV-overlapping probes has implications for determining underlying mechanisms of disorders such as schizophrenia, which have thus far been elusive. To accelerate progress in understanding epigenetic epidemiology of human disease, it will be crucial for the field to either augment the Illumina platform to improve coverage of CpG sites within CoRSIVs or adopt a next-generation standardized platform for profiling CoRSIV methylation in humans. Our results suggest that for many of the thousands of EWAS that have employed the Illumina platform it will be hugely advantageous to utilize the same DNA samples to perform methylation profiling at CoRSIVs. This presents an outstanding opportunity to rapidly advance our understanding of epigenetic etiology of disease. Lastly, the current list of known CoRSIVs is incomplete. The largest published screen for CoRSIVs (*27*) was based on only ten White American GTEx donors. To broadly explore associations between interindividual variation and risk of disease, it will be crucial to map out CoRSIVs across diverse human ancestry groups and environmental contexts.

## Data Availability

All data are available in the main text or the supplementary materials.

## Author contributions

Conceptualization: RAW

Methodology: WC, RAW

Data curation: WC

Formal analysis: WC, UM, AY

Funding acquisition: RAW

Supervision: CJG, RAW

Writing – original draft: WC, RAW

Writing – review & editing: all authors

## Data and materials availability

All data are available in the main text or the supplementary materials. All codes and pipelines to analyze data and generate figures are available on GitHub (https://github.com/computational-epigenetics-section/CoRSIV-text-mining).

## Supplementary Material

Materials and Methods

Figs. S1 to S15

Tables S1 to S22

References (54-58)

