## Supplementary material for "Human genomic regions of systemic interindividual epigenetic variation are implicated in neurodevelopmental and metabolic disorders": Materials and Methods, Supplementary Figures S1-S15

**The PDF file includes:**

Materials and Methods
Figs. S1 to S15
References (*54*-*58*)

**Other Supplementary Materials for this manuscript include the following:**

Tables S1 to S22

### Materials and Methods

Identification of EWAS reporting Illumina probe IDs

Medical Subject Headings (MeSH) comprise a defined, hierarchically organized vocabulary used in indexing articles in PubMed. Based on disease categories in a previous study (*28*), and the MeSH hierarchy under "diseases" [MeSH], we selected 11 disease categories for analysis: cancer, cardiovascular, digestive, endocrine, hematological, immune, metabolic, neurological (including both nervous system and mental disorders), obesity, respiratory, and urogenital (Fig. 1A).

We performed our search on January 16th, 2024; studies published thereafter are not included. Our analysis includes only studies listed in PubMed Central, an archive of free full-text biomedical literature. Supplementary files were available only if they were in the PubMed Central Open Access Subset and were accessed in csv and excel formats only. Employing NCBI's Entrez Programming Utilities, we used the following PubMed search terms: ("DNA Methylation"[MeSH Terms] AND "disease-specific term" [MeSH Terms]) NOT ("animals"[MeSH Terms] NOT "humans"[MeSH Terms]) AND "pubmed pmc"[Filter], retrieving 15,884 full-text studies (Fig. 1B).This double-negative syntax effectively excludes all studies containing only animal data and no human data limits the results to human studies by first identifying animal-only studies then excluding them (*54*).

Approximately 85% of published EWAS have utilized Illumina methylation arrays (*55*). Accordingly, and to exploit the uniform formatting of Illumina probe IDs, we limited our analyses to studies performed with the Illumina platform. After retrieving the full-text data set, we filtered to include only studies (N = 4,195) mentioning either at least one Illumina-related keyword ("450K", "HM450", "EPIC", "450k", "HumanMethylation450", "850K", "850k") or any Illumina probe ID (format: "cg" directly followed by 8 numeric digits). We then batch-downloaded any supplementary files for these 4,195 studies from the PubMed Central Open Access Subset. Of these, only 2,343 contained any Illumina probe IDs in either the main text or supplementary tables. As a final step, to limit our analysis to primary research articles only we excluded publications with PubMed article type tags including Editorial, Letter, News, Dataset, Meta-Analysis, Review, Systematic Review, Retracted Publication, Comment, Video-Audio Media, Validation Study, or Historical Article, leaving 2,203 studies in the data set (Fig 1B). Whereas most of these reported fewer than 1,000 Illumina probe IDs (fig. S1), some included supplementary tables listing hundreds of thousands of probes. We therefore excluded all supplementary tables reporting more than 1,000 probe IDs.

CoRSIVs and control regions used in this analysis

We compiled curated systemic interindividual variation (SIV) loci from several sources including putative metastable epialleles (*31*), Illumina probes identified as exhibiting epigenetic supersimilarity or systemic interindividual variation (*32*), and the 9,926 human CoRSIVs (*27*). Although some of these loci were identified prior to introduction of the CoRSIV terminology, we consider them all CoRSIVs. Of these 10,388 regions, only 1,607 (15.5%) are covered by at least one probe on the Illumina HM450 or EPIC array (table S1). Within these 1,607 regions are 3,517 probes, which we term CoRSIV probes (average 2.2 probes/CoRSIV); table S2 annotates these relative to proximal genes.

To establish a stable baseline from which to evaluate enrichment, we selected 10 sets of control regions from the human genome (table S3, fig. S2); within each set, one randomly selected control region was individually matched to each of the 10,388 CoRSIVs by (in decreasing priority): chromosome, region size, number of Illumina HM450 / EPIC probes, number of CpG sites, and proximity to genic features (number of genic transcription start sites, gene bodies, and transcription end sites within ± 3kb of region, respectively). Exact matching was required for chromosome, region size, and number of overlapping Illumina probes. This yielded 35,170 control probes (table S4) within 16,070 control regions. We averaged across the ten sets of control regions to establish a stable baseline from which to evaluate enrichment.

Gene set enrichment analysis and genic feature annotation

Gene annotations of Illumina probes were obtained from the "Unique UCSC RefGene Name" column of the official Illumina manifest. Gene set enrichment analyses were performed using the DisGeNET (*56*) library in Enrichr (*57*). Genic feature annotations of Illumina probes were based on the "UCSC_RefGene_Group" column of the official Illumina manifest.

Enrichment Ratio Calculation

Relative enrichment of CoRSIV probes within each MeSH disease category was calculated as the ratio of the weighted sum of CoRSIV probe occurrences to the average weighted sum of control probe occurrences, with weights based on the number of publications reporting each probe:

$$Enrichment = \frac{CoRSIV\ probe\ occurrences}{Control\ probe\ occurrences} = \frac{\sum_{i=1}^{k} i \cdot \frac{n_i}{p_i}}{\sum_{i=1}^{k} i \cdot \frac{m_i \div 10}{p_i}}$$

where $k$ is the highest number of papers that each report at least 10 identical probe IDs, $n_i$ is the number of CoRSIV probes reported in exactly $i$ papers, $p_i$ is the total number of probes reported in exactly $i$ papers, and mi is the number of control probes reported in exactly $i$ papers (across 10 control sets). We then visualized the MeSH hierarchy for each enriched category using Graphviz, omitting duplicated MeSH terms for clarity. To further simplify the MeSH hierarchies, we displayed only disease nodes representing at least 15 papers. Due to its larger size, we limited the cancer hierarchy to disease nodes representing at least 20 papers.

Permutation testing to establish statistical significance of enrichments

Within each disease category, we determined the null distribution by performing 100,000 iterations. Within each iteration, we first randomly permuted (shuffled) "CoRSIV" and "Control" labels to model the null hypothesis, then determined the permuted "CoRSIV" enrichment based on the enrichment formula above. Based on the normal distribution of the 100,000 permuted ratios, we then calculated the z-score and p value for the actual enrichment ratio. A disease category was considered enriched if it passed all three following criteria: (i) enrichment ratio > 1, (ii) permutation testing p-value remains significant after adjustment for multiple testing, and (iii) any of the CoRSIV probes driving the enrichment are reported in at least two papers.

Additional datasets to evaluate temporal stability, interindividual variation, and blood-brain concordance

We analyzed data from Flanagan et al. on long-term stability of DNA methylation in peripheral blood of 92 women ranging from 35 to 84 years old at baseline who were profiled twice using the Illumina HM450 methylation array, across an average follow-up period of six years (*35*) (GEO accession: GSE61151). To quantify long-term stability of probe-level methylation, we calculated the intraclass correlation coefficient (ICC) using a two-way random-effects model for single measures (*58*). And, as described previously (*36*), for each probe we calculated the 2nd to the 98th percentile of interindividual range ($IIR_{2-98\%}$) to provide a reliable estimate of interindividual variation. To examine brain-blood methylation concordance, we queried the BECon database (*37*), which provides probe-level data on methylation correlation between blood and four different brain regions.

Sensitivity Analysis

For each of the 11 MeSH categories, we performed a sensitivity analysis on probe-level ICC and $IIR_{2-98\%}$ to assess these in relation to the number of papers in which CoRSIV and control probes were reported. Specifically, we analyzed median ICC vs. median $IIR_{2-98\%}$ values for CoRSIV and control probes across subsets of probes reported in increasing numbers of papers. Within each of the 11 MeSH categories, we also performed sensitivity analyses on probe-level BECon scores (*37*) to determine if the brain-blood correlation for CoRSIV and control probes is associated with the number of papers reporting the probe.

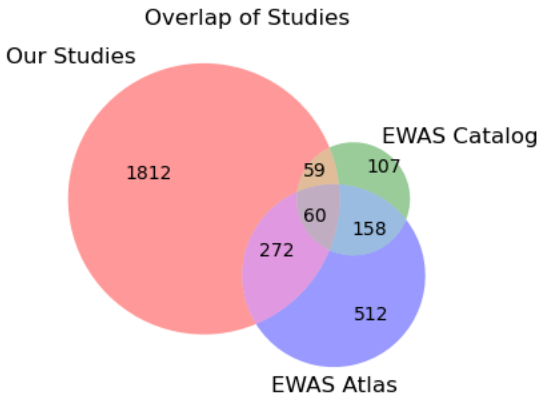


**Fig. S1.**
**Overlap of studies across the set we curated, the EWAS Catalog, and the EWAS Atlas.**

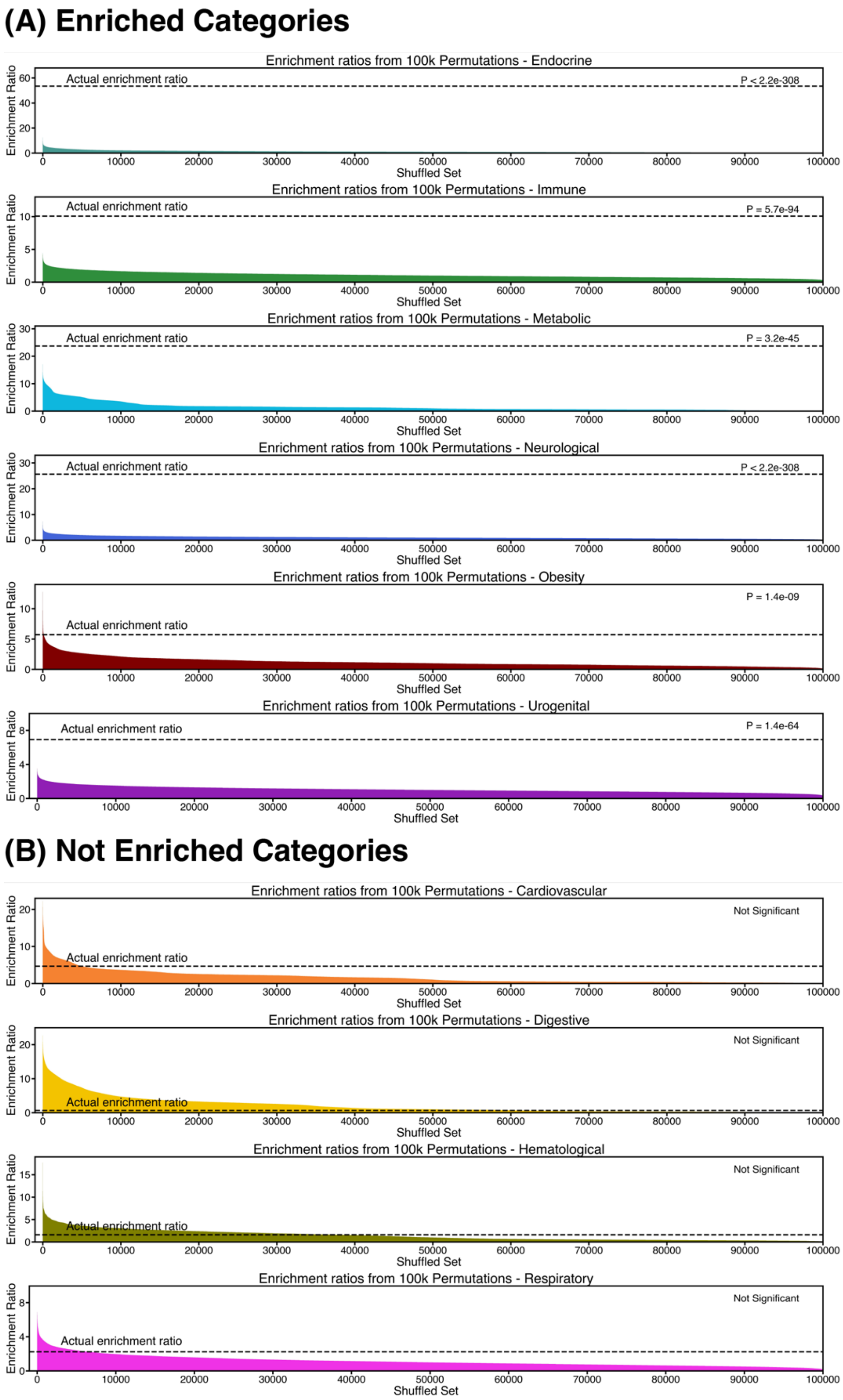


**Fig. S2.**

**Results of permutation testing (100,000 iterations) for each disease category.** Enriched categories are displayed in (**A**), and not-enriched categories in (**B**).

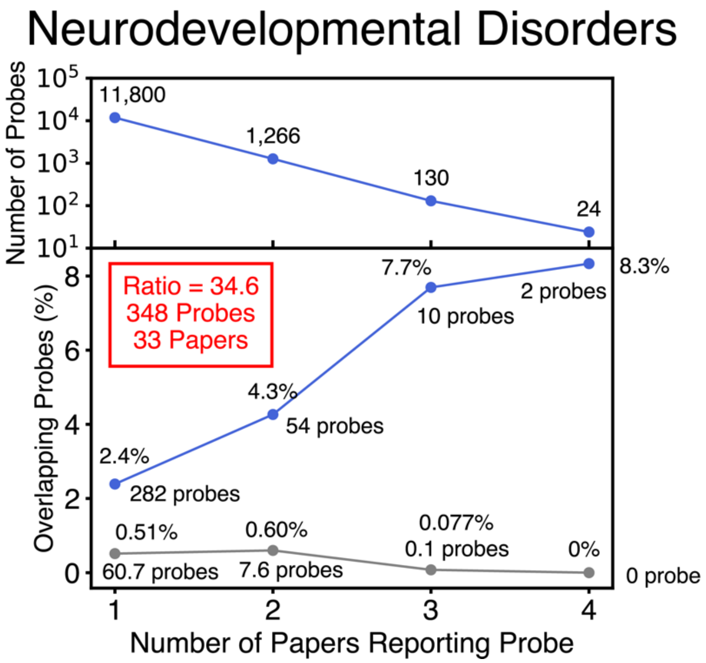


$$\text{Enrichment} = \frac{\sum_{i=1}^{k} i \cdot \frac{n_i}{p_i}}{\sum_{i=1}^{k} i \cdot \frac{m_i \div 10}{p_i}}$$

$$= \frac{1 \cdot \frac{n_1}{p_1} + 2 \cdot \frac{n_2}{p_2} + 3 \cdot \frac{n_3}{p_3} + 4 \cdot \frac{n_4}{p_4}}{1 \cdot \frac{m_1 \div 10}{p_1} + 2 \cdot \frac{m_2 \div 10}{p_2} + 3 \cdot \frac{m_3 \div 10}{p_3} + 4 \cdot \frac{m_4 \div 10}{p_4}}$$

$$= \frac{1 \times \frac{282}{11800} + 2 \times \frac{54}{1266} + 3 \times \frac{10}{130} + 4 \times \frac{2}{24}}{1 \times \frac{60.7}{11800} + 2 \times \frac{7.6}{1266} + 3 \times \frac{0.1}{130} + 4 \times \frac{0}{24}}$$

$$= \frac{1 \times 2.4\% + 2 \times 4.3\% + 3 \times 7.7\% + 4 \times 8.3\%}{1 \times 0.51\% + 2 \times 0.60\% + 3 \times 0.077\% + 4 \times 0\%} = \frac{67.3}{1.94} = 34.6$$

**Fig. S3.**

**Annotated example illustrating calculation of the enrichment ratio for the neurodevelopmental disorders category.** The strong enrichment is largely driven by the 12 CoRSIV probes that are each reported in 3-4 papers.

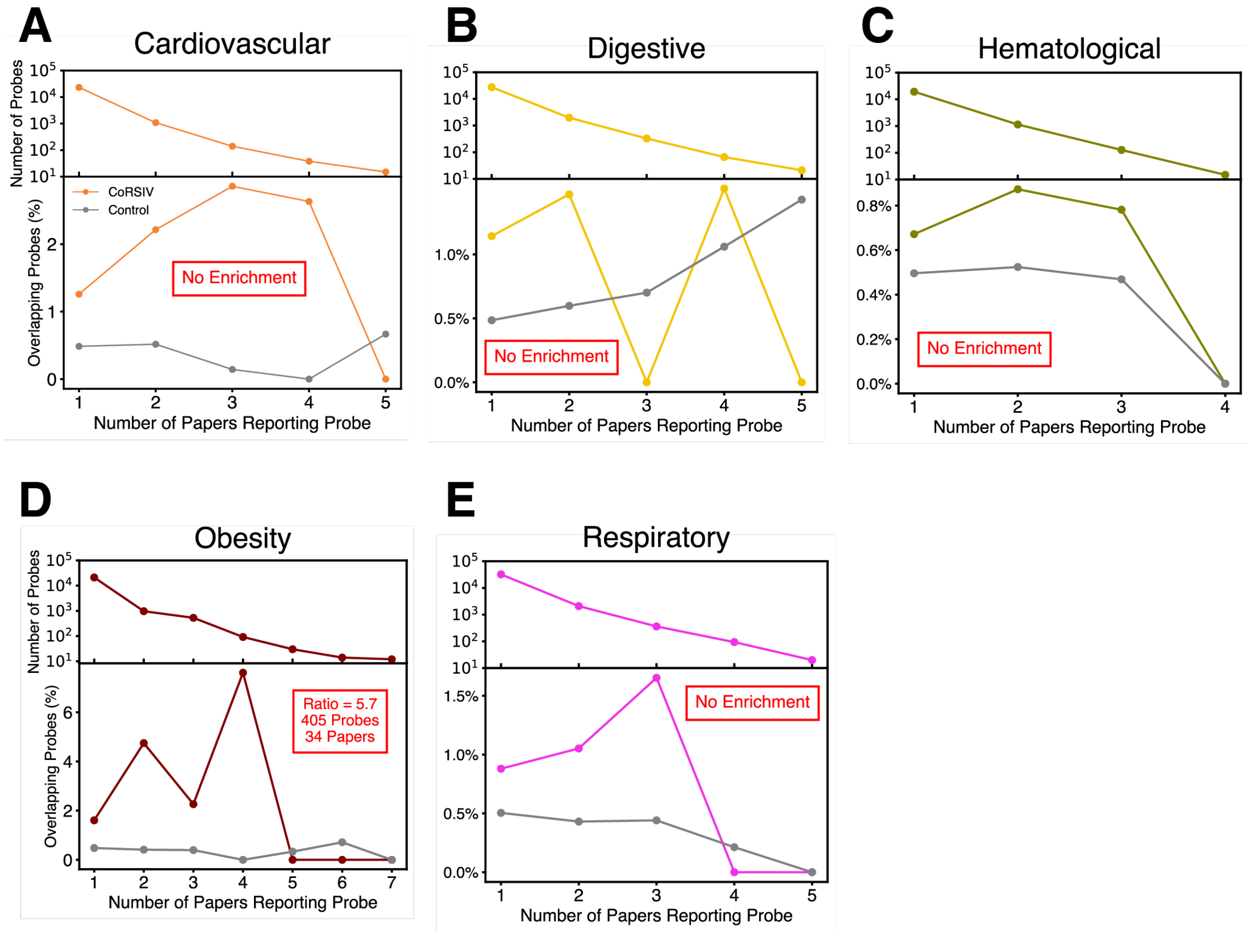


**Fig. S4.**

**Results for five additional categories.** Stacked decay and enrichment plots for (**A**) cardiovascular, (**B**) digestive, (**C**) hematological, (**D**) obesity, and (**E**) respiratory categories, respectively. Of these, only obesity shows enrichment.

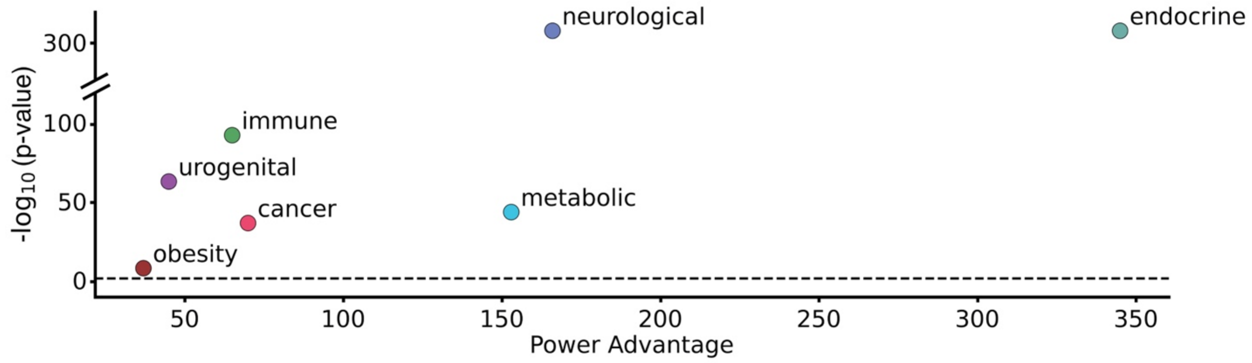


**Fig. S5.**

**Power advantage within enriched disease categories.** This illustrates the expected power advantages of EWAS profiling all ~10,000 known CoRSIVs, relative to using the EPIC array.

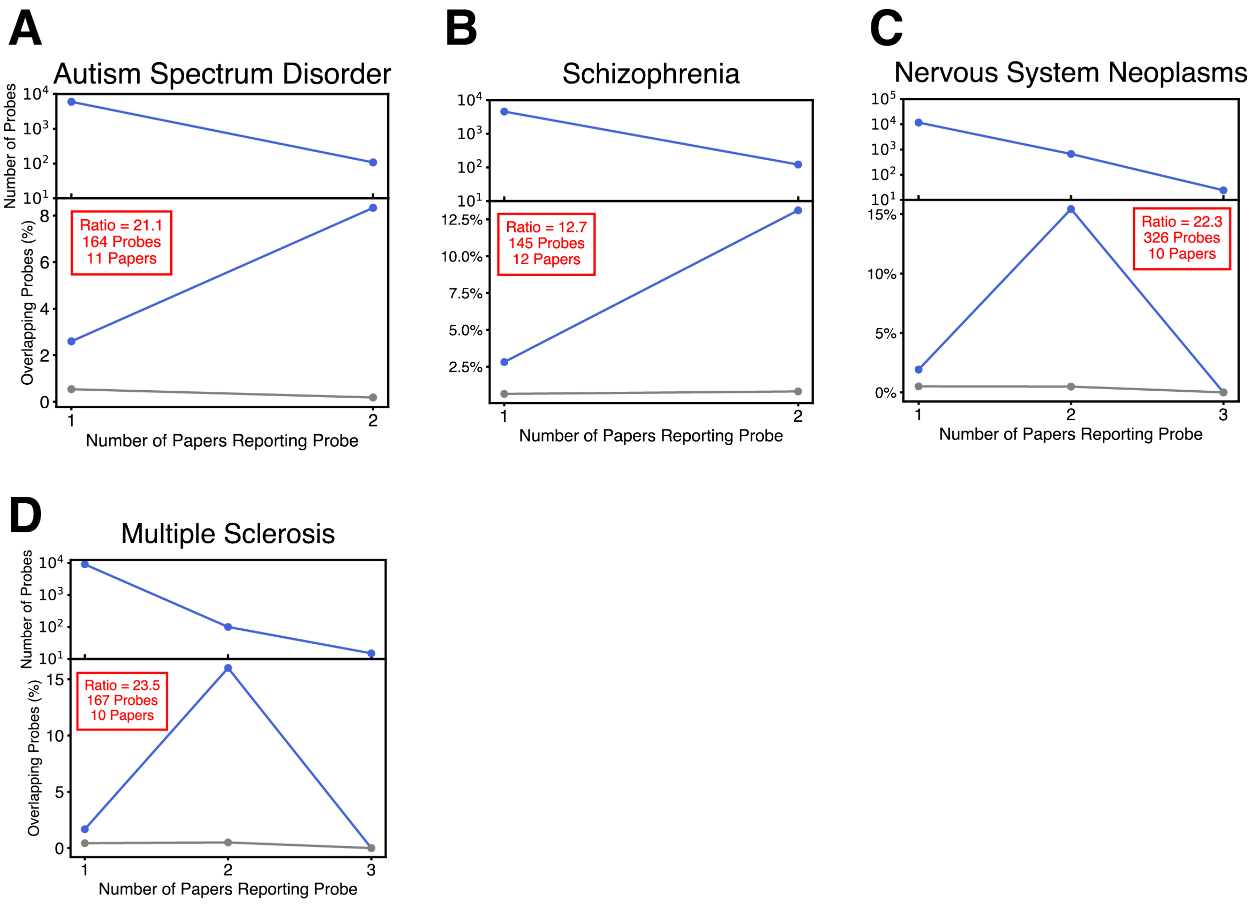


**Fig. S6.**

**Stacked decay and enrichment plots for additional enriched disorders within the neurological category.** (**A**) Autism spectrum disorder ($P = 4.0 \times 10^{-108}$), (**B**) Schizophrenia ($P = 1.0 \times 10^{-97}$), (**C**) Nervous system neoplasms ($P < 2.2 \times 10^{-308}$), (**D**) Multiple sclerosis ($P = 8.4 \times 10^{-291}$). Insets (red boxes) indicate enrichment ratio and the number of probes and publications driving each enrichment.

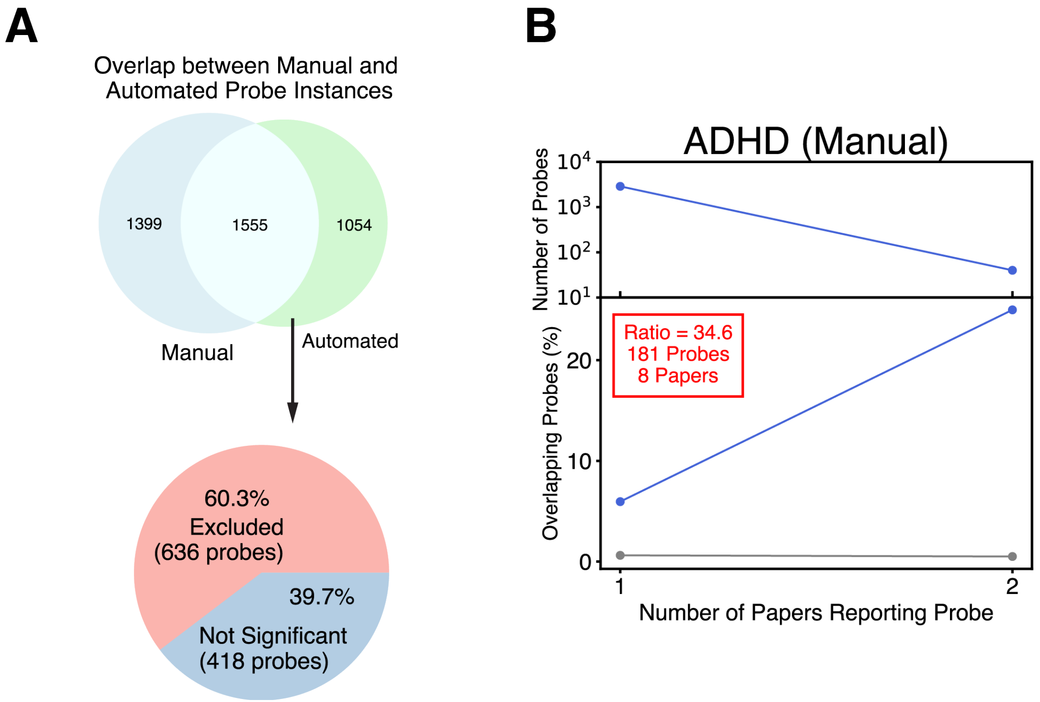


**Fig. S7.**

**A manually curated analysis of ADHD yields results comparable to the automated approach**. (**A**) Venn diagram showing the overlap of probe instances collected from the manual and automated approaches (top) and pie chart categorizing why 1,054 probe instances were found only in the automated approach. 636 probes were excluded from the manual approach because they were not directly relevant to ADHD outcomes, primarily consisting of probes for which methylation differences resulted from exposures. (**B**) Stacked decay and enrichment plots obtained in the manually curated analysis of ADHD based on nominally significant probes ($P<0.05$). The enrichment ratio is comparable to the one we obtained from our automated approach (54.7-fold) and is statistically significant by permutation testing ($P = 3.0 \times 10^{-276}$). Inset (red box) indicates enrichment ratio and the number of probes and publications driving each enrichment.

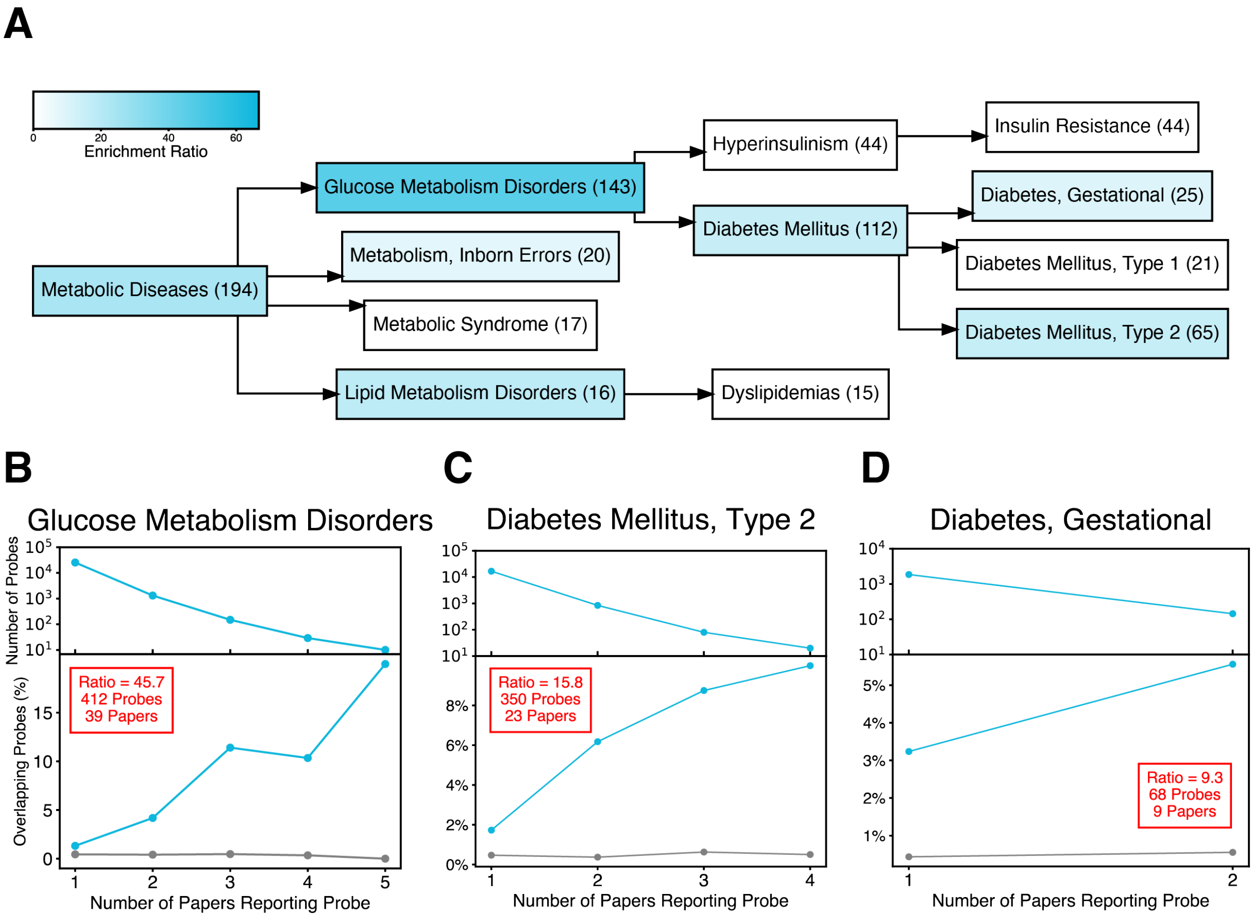


**Fig. S8.**

**CoRSIVs are associated with type 2 but not type 1 diabetes.** (**A**) Term-specific results for the MeSH hierarchy encompassing the metabolic category. Statistically significant enrichments are indicated by teal shading. Numbers indicate the total number of papers analyzed within each MeSH term. Only categories showing (i) statistically significant enrichment, and (ii) including at least one CoRSIV probe reported in at least two papers are colored. (**B-D**) Stacked decay and enrichment plots for glucose metabolism disorders ($P = 7.8 \times 10^{-170}$), type 2 diabetes ($P = 2.0 \times 10^{-32}$), and gestational diabetes ($P = 1.6 \times 10^{-29}$), respectively. Insets (red boxes) indicate enrichment ratio and the number of probes and publications driving each enrichment.

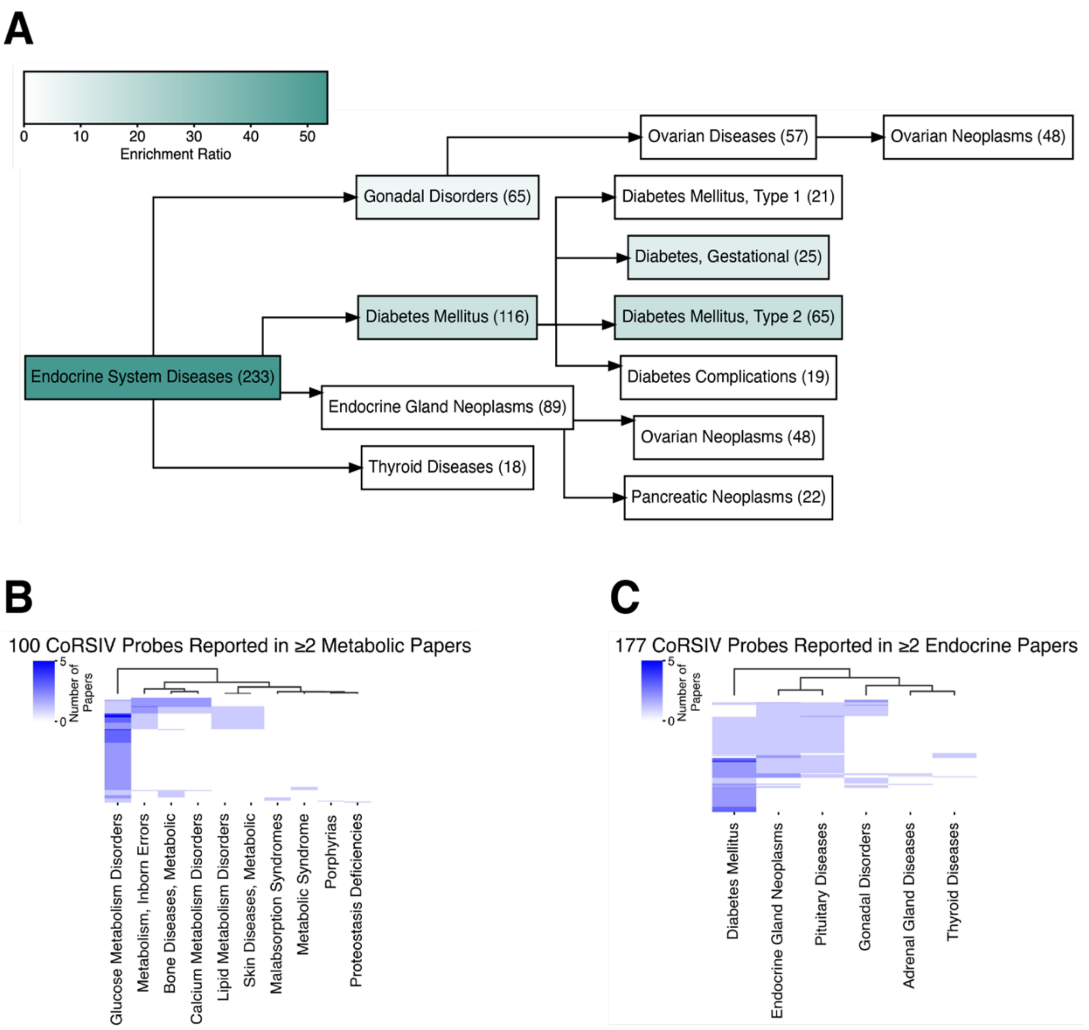


**Fig. S9.**

**The strong enrichment for the endocrine category is driven by many of the same probes being reported in multiple sub-categories.** (**A**) Term-specific results for the MeSH hierarchy encompassing the endocrine category. Statistically significant enrichments are indicated by green shading. Numbers indicate the total number of papers analyzed within each MeSH term. Only categories showing (i) statistically significant enrichment, and (ii) including at least one CoRSIV probe reported in at least two papers are colored. (**B**) Unsupervised hierarchical clustering of 100 CoRSIV probes reported in two or more studies in the metabolic category reveals that most probes are not shared across multiple sub-categories. (**C**) Conversely, unsupervised hierarchical clustering of 177 CoRSIV probes reported in two or more studies in the endocrine category shows substantial overlap of probes across multiple sub-categories, explaining the overall high enrichment of the "Endocrine System Diseases" MeSH heading. Note that “Pituitary Diseases” and “Adrenal Gland Diseases” were omitted in panel A because they contain less than 15 papers.

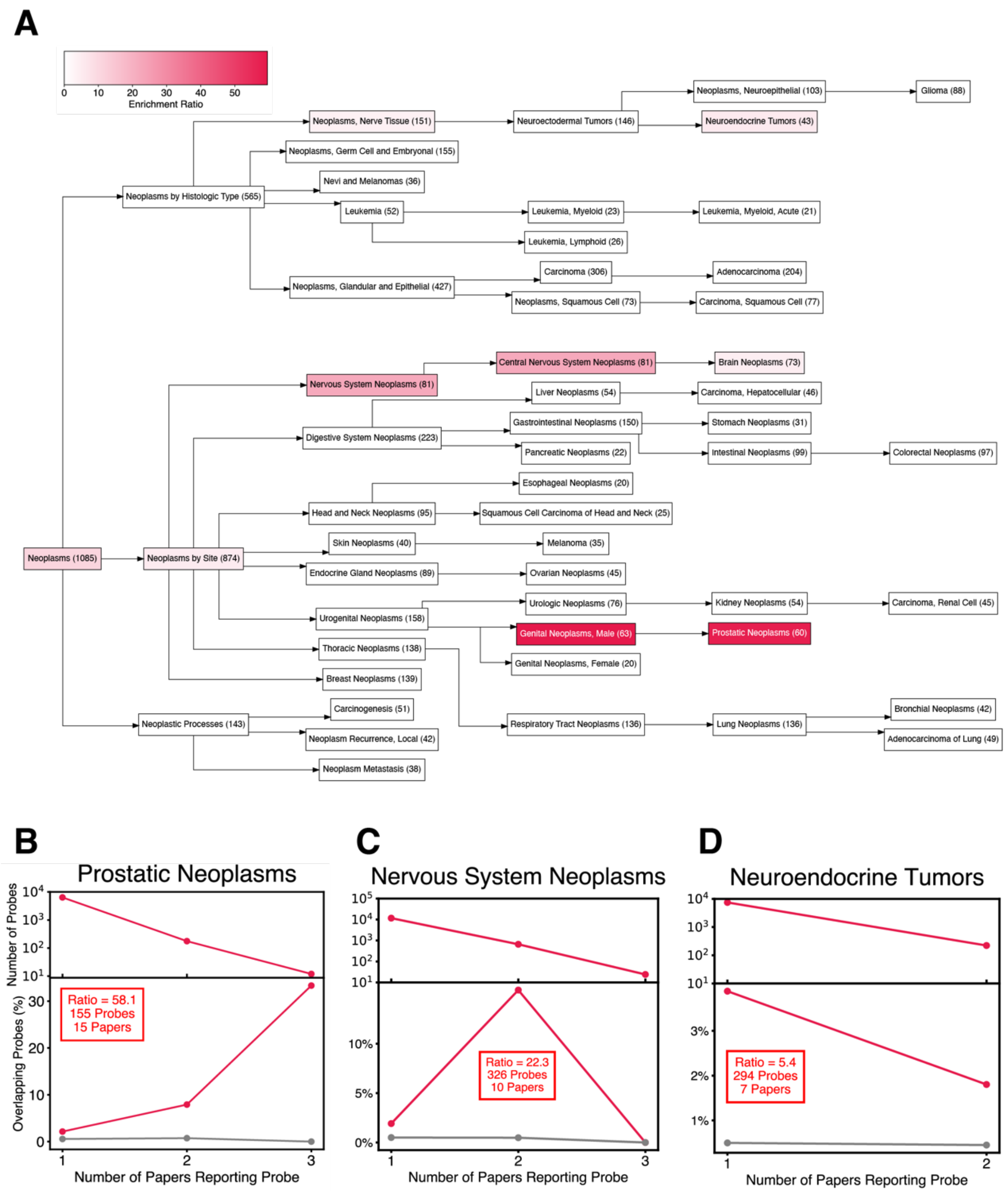


**Fig. S10.**

**CoRSIVs are associated with prostate cancer.** (**A**) Term-specific results for the MeSH hierarchy encompassing the cancer category. Statistically significant enrichments are indicated by red shading. Numbers indicate the total number of papers analyzed within each MeSH term. Only categories showing (i) statistically significant enrichment, and (ii) including at least one CoRSIV probe reported in at least two papers are colored. (**B-D**) Stacked decay and enrichment plots for prostate cancer ($P = 1.2 \times 10^{-87}$), nervous system neoplasms ($P < 2.2 \times 10^{-308}$) and neuroendocrine tumors ($P < 2.7 \times 10^{-14}$). Insets (red boxes) indicate enrichment ratio and the number of probes and publications driving each enrichment.

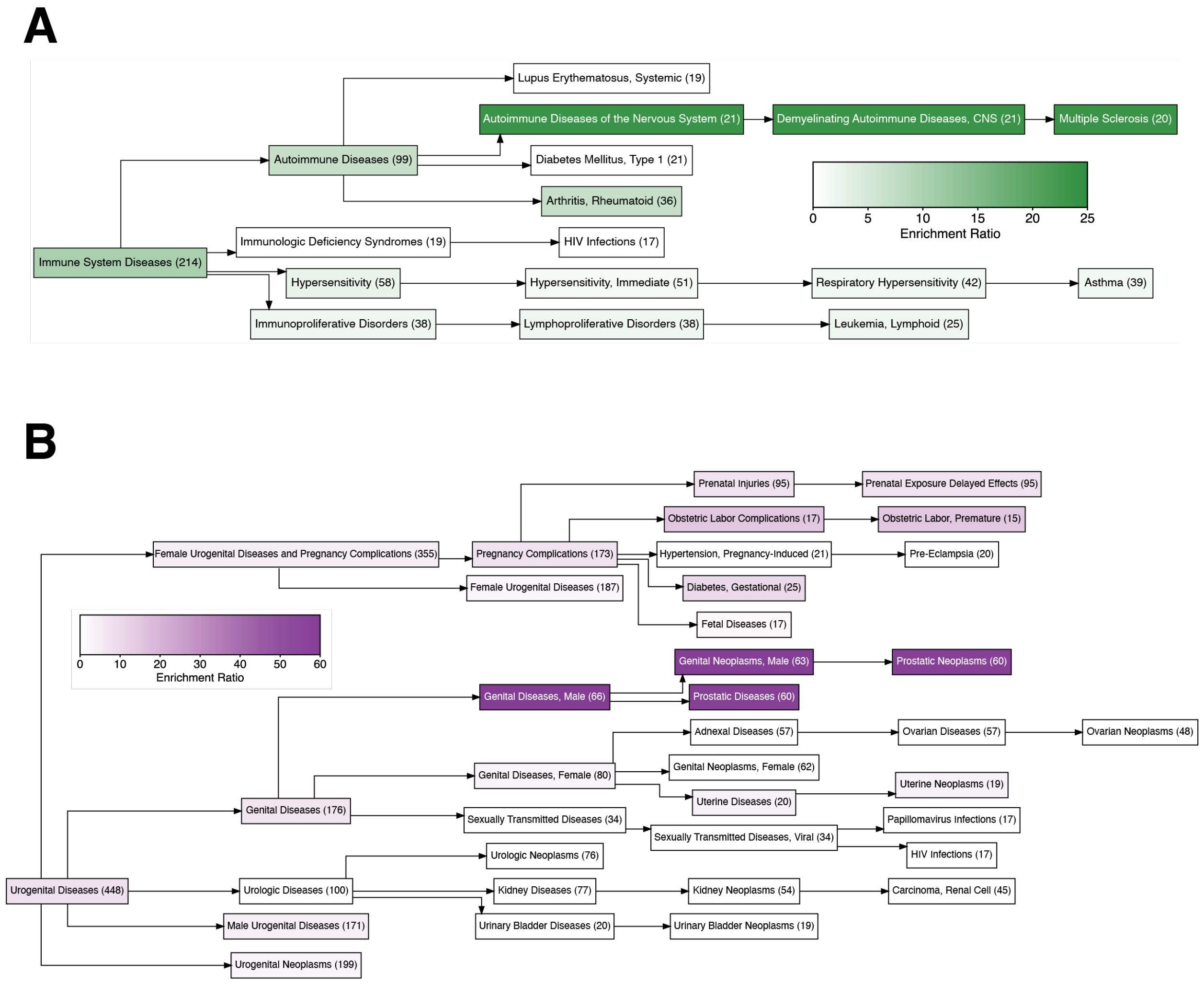


**Fig. S11.**

**Enrichment results for the immune and urogenital MeSH Trees.** Term-specific results for the MeSH hierarchy encompassing the (**A**) immune and (**B**) urogenital categories. Statistically significant enrichments are indicated by green and purple shading, respectively. Numbers indicate the total number of papers analyzed within each MeSH term. Only categories showing (i) statistically significant enrichment, and (ii) including at least one CoRSIV probe reported in at least two papers are colored.

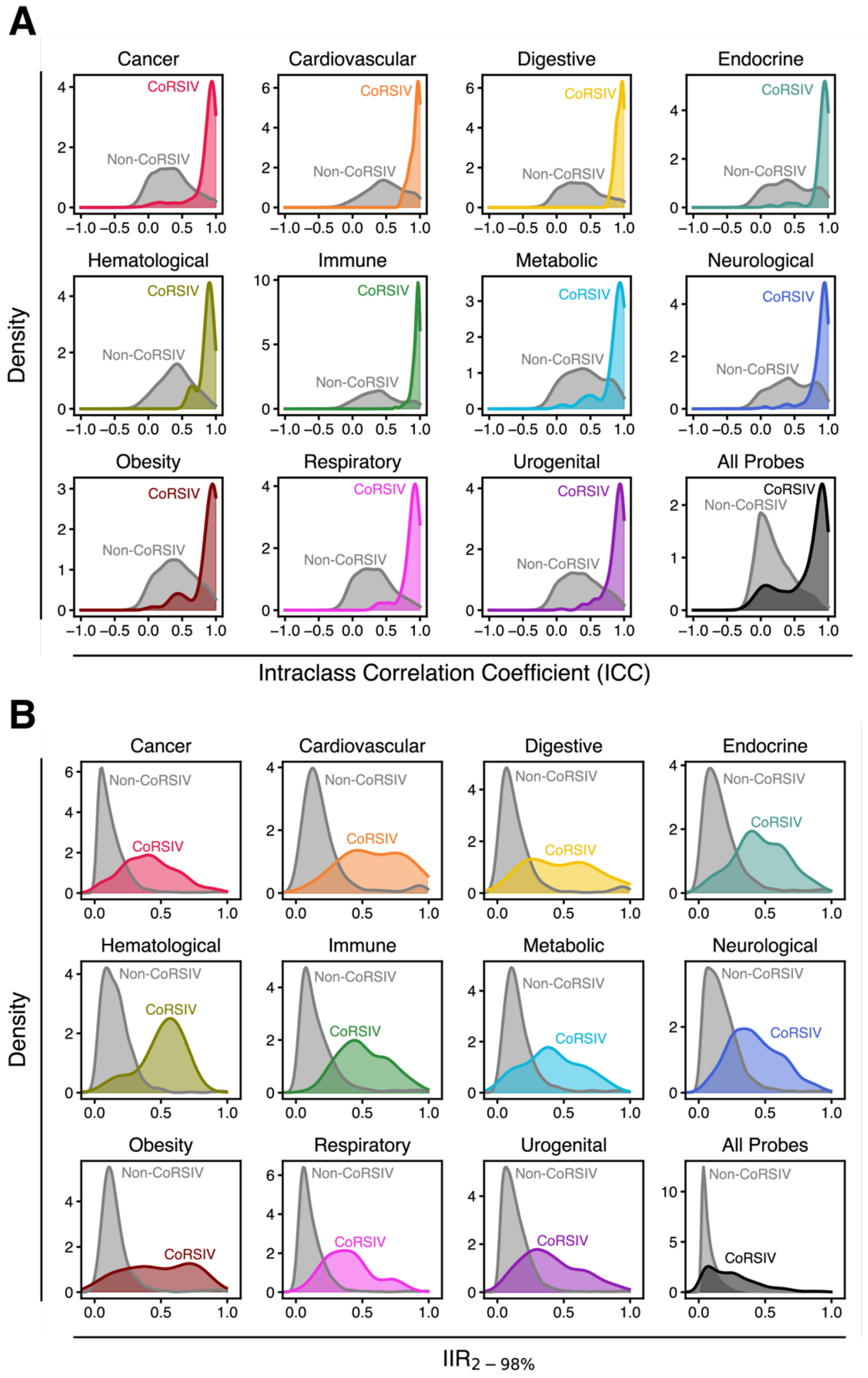


**Fig. S12.**

**Within all categories, stability and interindividual variation of CoRSIV probes are greater than those of non-CoRSIV probes.** (**A**) ICC and (**B**) $IIR_{2\text{-}98\%}$ distribution for probes reported in at least two papers, as well as all CoRSIV vs Non-CoRSIV probes. CoRSIV probe distributions are colored.

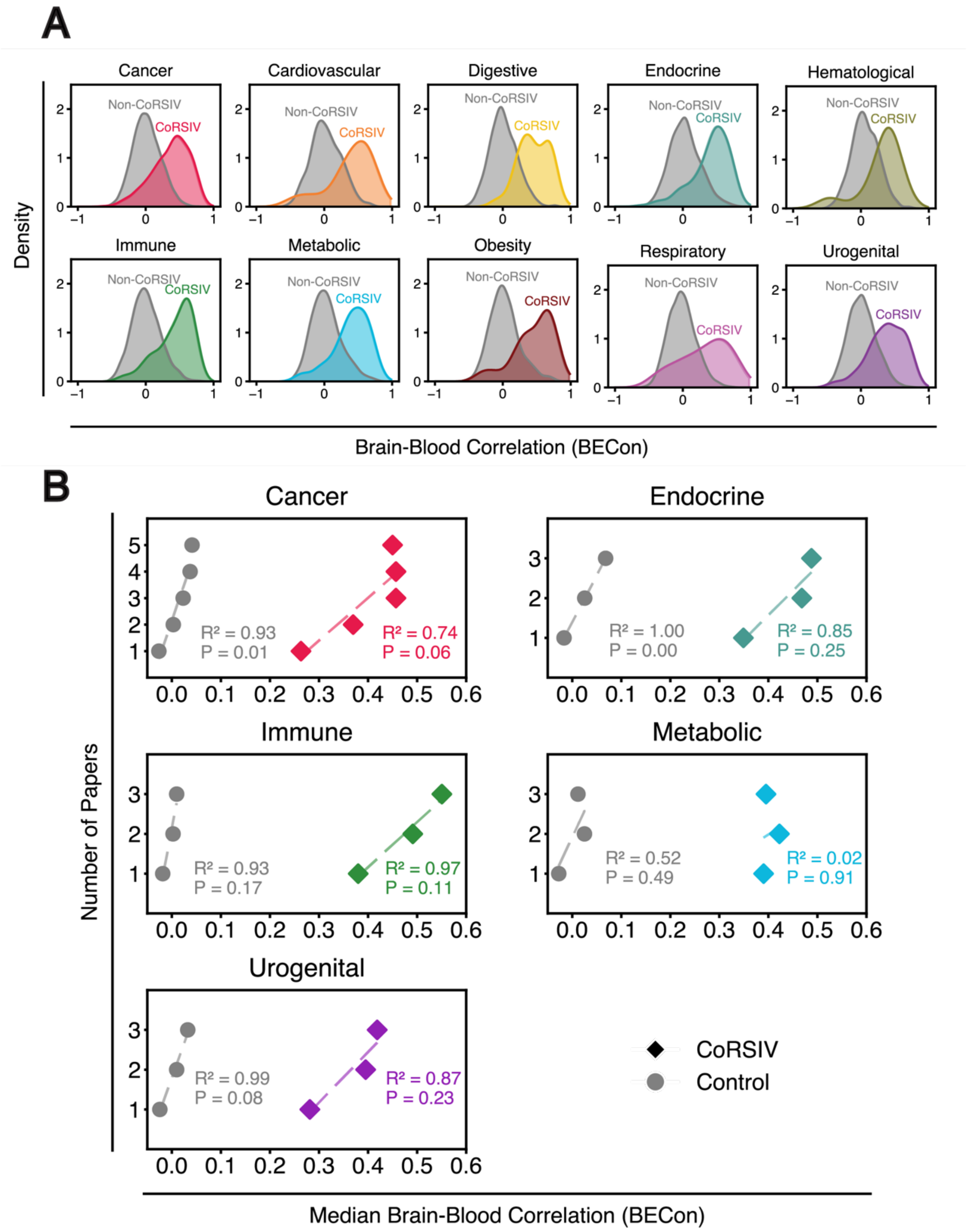


**Fig. S13.**

**Results of BECon analysis for categories other than neurological.** (**A**) BECon score distribution for probes reported in at least two papers. Within each category, CoRSIV probes show higher BECon scores than non-CoRSIV probes. (**B**) Sensitivity analyses. Unlike in the neurological category (Fig. 5C), being reported in more cancer, endocrine, immune, metabolic, or urogenital papers is not associated with increasing BECon score for CoRSIV probes.

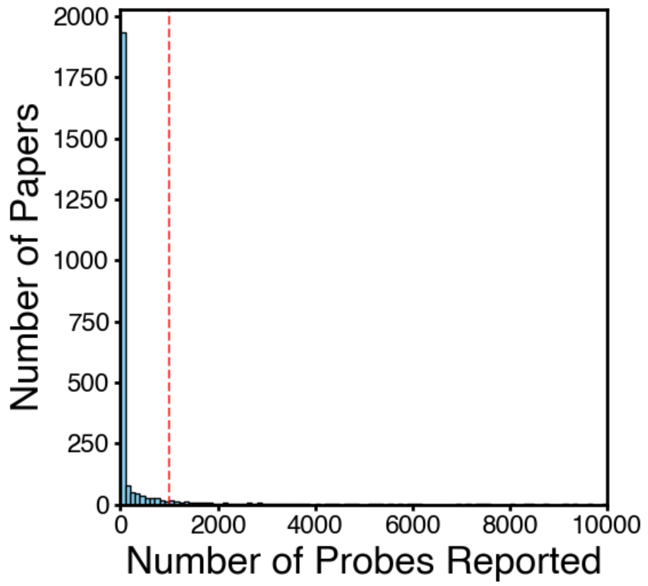


**Fig. S14.**

**Cutoff for Maximum Number of Probes per Study.** Distribution of 2,203 studies based on the total number of probes reported within the main text and supplementary tables. Most papers report fewer than 1,000 probes (red dotted line).

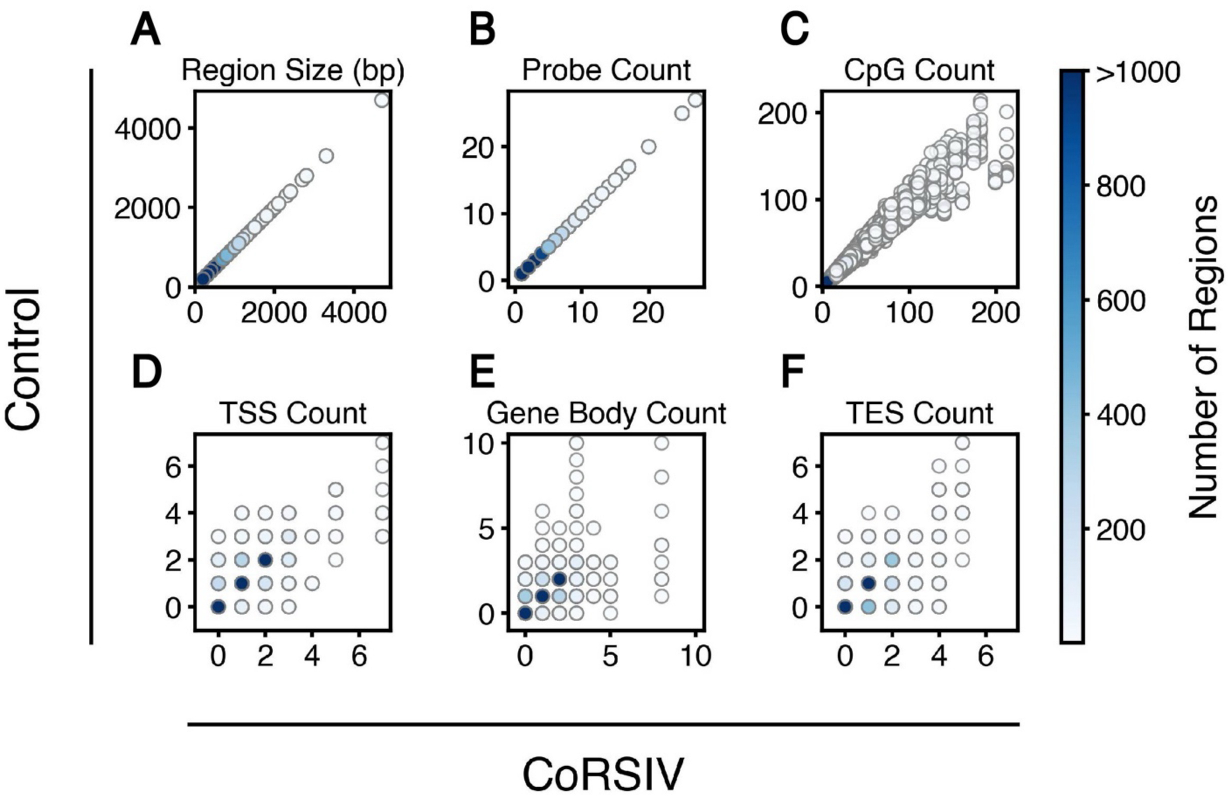


**Fig. S15.**

**CoRSIV vs. Control Region characteristics.** In addition to matching by chromosome, control regions were matched with CoRSIVs on the following characteristics: perfectly on (**A**) region size and (**B**) number of Illumina probes, and closely on (**C**) number of CpGs, (**D**) number of transcription start sites (TSS) within ±3kb, (**E**) number of gene bodies within ±3kb, and (**F**) number of transcription end sites (TES) within ±3kb. Each point represents one of the 16,070 control regions. The color scale is capped at 1,000 regions.

**Supplementary Table Legends.**

**Table S1.** Information on all 2,203 studies included in this study.

**Table S2.** 41,653 probes were reported in $\geqslant$ 2 papers.

**Table S3.** Significant (Adjusted-P < 0.05) DisGeNET results from Enrichr, based on probes reported in $\geqslant$2 papers only under each category.

**Table S4.** Annotations on 1,607 CoRSIVs covered on the HM450 / EPIC array.

**Table S5.** Annotations on all 3,517 CoRSIV probes covered on the HM450 / EPIC array.

**Table S6.** Annotations on all 35,170 control HM450 / EPIC probes.

**Table S7.** Annotations on all 16,070 controls (10 sets for each CoRSIV).

**Table S8.** Enrichment ratios and permutation testing P values for each of the 7 statistically significantly enriched main categories.

**Table S9.** Enrichment ratios and permutation testing P values for significantly enriched diseases under neurological category.

**Table S10.** Nominally significant probes from ADHD studies.

**Table S11.** Enrichment ratios and permutation testing P values for significantly enriched disease under metabolic category.

**Table S12.** Enrichment ratios and permutation testing P values for significantly enriched disease under endocrine category.

**Table S13.** Enrichment ratios and permutation testing P values for significantly enriched disease under cancer category.

**Table S14.** Enrichment ratios and permutation testing P values for significantly enriched disease under immune category.

**Table S15.** Enrichment ratios and permutation testing P values for significantly enriched disease under urogenital category.

**Table S16.** List of cancer papers reporting CoRSIV probes that appear in at least two cancer studies.

**Table S17.** List of endocrine papers reporting CoRSIV probes that appear in at least two endocrine studies.

**Table S18.** List of immune papers reporting CoRSIV probes that appear in at least two immune studies.

**Table S19.** List of metabolic papers reporting CoRSIV probes that appear in at least two metabolic studies.

**Table S20.** List of neurological papers reporting CoRSIV probes that appear in at least two neurological studies.

**Table S21.** List of urogenital papers reporting CoRSIV probes that appear in at least two urogenital studies.

**Table S22.** $IIR_{2\text{-}98\%}$ and ICC for Flanagan data for all probes.
